# Amygdala subdivisions exhibit aberrant whole-brain functional connectivity in relation to stress intolerance and psychotic symptoms in 22q11.2DS

**DOI:** 10.1101/2022.09.02.22279263

**Authors:** Farnaz Delavari, Halima Rafi, Corrado Sandini, Ryan Murray, Caren Latrech, Dimitri Van De Ville, Stephan Eliez

## Abstract

The amygdala is a key region in emotional regulation, which is often impaired in psychosis. However, it is unclear if amygdala dysfunction directly contributes to psychosis, or whether it contributes to psychosis through symptoms of emotional dysregulation. We studied the functional connectivity of amygdala subdivisions in patients with 22q11.2DS, a known genetic model for psychosis susceptibility. We investigated how dysmaturation of each subdivision’s connectivity contributes to positive psychotic symptoms and impaired tolerance to stress in deletion carriers.

Longitudinally-repeated MRI scans from 105 patients with 22q11.2DS (64 at high-risk for psychosis and 37 with impaired tolerance to stress) and 120 healthy controls between the ages of 5 to 30 years were included. We calculated seed-based whole-brain functional connectivity for amygdalar subdivisions and employed a longitudinal multivariate approach to evaluate the developmental trajectory of functional connectivity across groups.

Patients with 22q11.2DS presented a multivariate pattern of decreased basolateral amygdala (BLA)-frontal connectivity alongside increased BLA-hippocampal connectivity. Moreover, associations between developmental drops in centro-medial amygdala (CMA)-frontal connectivity to both impaired tolerance to stress and positive psychotic symptoms in deletion carriers were detected. SFA hyperconnectivity to the striatum was revealed as a specific pattern arising in patients who develop mild to moderate positive psychotic symptoms.

Overall, CMA-frontal dysconnectivity was found as a mutual neurobiological substrate in both impaired tolerance to stress and psychosis, suggesting a role in prodromal dysregulation of emotions in psychosis. While BLA dysconnectivity was presented as an early finding in patients with 22q11.2DS, contributing to impaired tolerance to stress.

## Introduction

Schizophrenia is often conceptualized as a gradually unfolding process that results in psychosis (1). Defining psychosis as a developmental process has allowed clinicians to view preclinical manifestations as targets for early intervention and preventive measures. Thus, in hopes of gaining new insights into the pathophysiology of psychosis vulnerability, there has been a surge in literature characterizing prodromal stages of schizophrenia in high-risk populations. In this regard, among the earlier clinical features of prodromal stage are symptoms such as depression, distress, and anxiety symptoms (2-5), rather than psychosis-specific manifestations (6). Furthermore, various studies have reported that anxiety and emotional dysregulation are associated with poorer prognoses for high-risk patients (7, 8). Specifically, reduced tolerance to daily life stress has been represented a critical symptom that mediates the transition of patients to psychosis (9-13). Still, the neurobiological mechanisms underlying the link between stress and psychosis vulnerability remain poorly characterized.

The amygdala is known as a key brain structure in emotional regulation, in particular fear, anxiety, and response to stress (14-16). It has also been suggested that abnormal amygdala activation may play a direct role in provoking psychotic-like symptoms. For example, epileptic activities in the amygdala are known to provoke sensory hallucinations and affective symptoms such as anxiety and fear (17-19). Indeed, neuroimaging studies have identified alterations of the amygdala in psychotic disorders, both in terms of structure and activity (20-25). The majority of volumetric studies report a reduction in the amygdala volume in clinically high-risk patients as well as in patients with psychosis (26-30). Functional findings are relatively more diverse, but overall, resting-state dysconnectivity between the amygdala and frontal regions is reportedly found in patients with psychotic disorders (20, 31-34).

However, it remains unclear if there is a direct link between the amygdala and psychosis, or whether amygdala dysfunction mediates symptoms of anxiety and stress intolerance, which finally settles as symptoms of psychosis. In this regard, it is worth noting that most of previous research in psychosis has studied the amygdala as a whole, while it is known that the amygdala comprises heterogeneous nuclei that perform distinct physiological tasks (35, 36). Considering amygdala as one uniform region potentially masks the distinct functional contribution of each individual division.

Functional and histological studies have allowed the identification of three functionally distinct subdivisions in the amygdala. The basolateral group (BLA), often characterized as the “inlet” of the amygdala, receives connections from various cortical and subcortical regions and participates in emotional learning and fear processing (37, 38). The centro-medial group (CMA) or the “outlet” of the amygdala mainly projects to the brainstem and hypothalamus as well as the striatum and prefrontal cortex, forming the behavioral response to the received information (38, 39). The superficial group (SFA) connects to the orbitofrontal cortex and participates in social affective functioning and olfactory processes (38, 40). Notably, the distinct assortment of cortical and subcortical connections of each amygdala subdivision, also undergo different developmental trajectories from childhood to adulthood (41) (42). For instance, in the typically developing brain, distinctive connections from the CMA and BLA to subcortical regions have been found in childhood, whereas this dissociation does not appear for cortical connections until adulthood (43). The faster maturation of the amygdala-subcortical connections is indicative of bottom-up emotional regulation, which is needed in early life to respond to critical stimuli (44, 45). Through development, long-range connections to BLA form to support the amygdala’s function in emotional and fear learning (46, 47). As such, mature amygdala-frontal connections are known to form by late adolescence to allow for the cognitive regulation of emotions (48).

Thus, what is defined as normal function in the amygdala may differ in childhood, adolescence, and adulthood. In the context of psychosis, this is particularly important since adolescence is a critical period when prodromal symptoms usually arise (49). Therefore, investigating developmental deviations of each amygdalar subdivisions from their normal function is critical to gain insight into the underlying mechanisms of the prodromal stage. To our knowledge only one study has investigated developmental changes of functional connectivity in two of the three amygdala subdivisions, CMA and BLA, in relation to psychosis. In doing so, Jalbrzikowski et al. (2019) have identified abnormal development of CMA functional connectivity during early adolescence in patients with psychosis disorders (50). Additionally, another study on adults with schizophrenia has identified abnormal SFA and CMA functional connectivity in adults with psychosis(51). Regarding how amygdala dysfunction and stress interact in psychosis, findings in mice models for schizophrenia points to a direct link between stress behavior caused by BLA hyperactivation, that aggravates a cascade of neurobiological changes that result in psychosis (52, 53). Yet, in humans, how abnormal maturation of amygdala subdivisions could make some individuals more vulnerable to psychosis, through pathways of stress intolerance, has not been investigated.

To investigate developmental vulnerability to psychosis, studying individuals with 22q11 deletions syndrome (22q11.2DS) offers an exceptional opportunity (54). Patients affected with 22q11.2DS are at a higher risk of developing various psychopathologies such as anxiety disorders, attention deficit hyperactivity disorders and mood disorders (55). Most strikingly, deletion carriers present up to 40% risk of developing psychosis by adulthood (56). Moreover, this syndrome often gets diagnosed due to somatic manifestations long before the neuropsychiatric symptoms emerge. 22q11.2DS has thus become a validated model for the longitudinal study of pre-clinical psychosis and investigation of neural biomarkers underlying psychopathology (55). Similar to schizophrenia, psychopathological manifestations in 22q11.2DS are not isolated and happen with high comorbidity rate (57). Moreover, evidence suggests that the presence of mood and anxiety disorder in patients with 22q11.2DS increases the risk of developing psychosis at follow-up (56, 58, 59).

Here, we aimed to characterize abnormal resting-state functional connectivity for all three functionally distinct amygdala subdivisions in patients with 22q11.2DS. We investigated how maturation patterns differ across deletion carriers who show mild to moderate positive psychotic symptoms as well as patients who manifest mild to moderate symptoms of impaired tolerance to stress. To do so, we employed a multivariate longitudinal design in a large cohort of patients with 22q11.2DS as well as in healthy controls across a wide age-span. We hypothesized that different amygdala subdivisions would show unique patterns of dysmaturation with distinctive association with psychopathology. In light of previous research, we expected the CMA to show a divergent developmental trajectory in patients at a higher risk for psychosis (PS) and contribute to symptoms of impaired tolerance to stress (ITS), and abnormal BLA connections to be more prominent in patients with ITS.

## Material and Methods

### Participants

The study population is part of the large prospective cohort on 22q11.2DS conducted in Geneva, Switzerland. Recruitment took place via the patients’ association and word of mouth. A total of 225 individuals were included in the study among which 105 participants (47.62% female) with 22q11.2DS were included and underwent consecutive longitudinal visits. Number of longitudinal assessments varied between 1 – 4, resulting in overall 160 longitudinal assessments. 22q11.2DS carriers were confirmed for diagnosis of 22q11.2DS thorough quantitative fluorescent polymerase chain reaction. Healthy controls (HCs; n= 120; 53.33% female) were enrolled from unaffected siblings of the patients or through an open call to the Geneva State School system in Switzerland. HCs were screened for history of any psychopathology and developmental abnormalities. Number of longitudinal visits varied between 1 - 3, resulting in overall 171 longitudinal assessments. In total 39.05% of patients and 35% of HC were followed by at least one other longitudinal visit. A detailed summary of repetitive assessments is provided in supplementary material. Both groups were matched for age, sex, number of scans and time between visits.

### Psychiatric Assessment

During each visit, 22q11 deletion carriers completed a comprehensive clinical assessment with an expert psychiatrist (SE). For 99 participants with the deletion this interview included the Structured Interview for Prodromal Syndromes(49, 60) which in line with previous literature on patients with 22q11D, was used to constitute subgroups of patients based on the participants’ clinical manifestation of positive psychotic symptoms (Unusual Thought Content, Suspiciousness, Grandiosity, Hallucinations, and Disorganized Communication)(61).Patients were considered PS(+) (positive symptoms; n =64) if they scored equal to or higher than 3 in at least one visit on any of the subscales for positive psychotic symptoms. The remaining participants were considered PS(−) (n=35). Similarly, patients who ever scored higher or equal to 3 in the subscale for impaired tolerance to normal stress were considered ITS(+) (impaired tolerance to stress; n=37) and the remainder of the patients were considered ITS(-)(n=62). In addition, a semi-structured clinical interview was conducted, to assess the presence of comorbid psychiatric conditions (62-64). Participants’ demographic and clinical characteristics are summarized in Table-1. Written informed consent was obtained from participants or their parents under protocols approved by the Swiss Ethical Committee (Commission Centrale d’Ethique de la Recherche, Geneva Canton).

|  | Number of scans | Number of subjects (F/M) | Age, Mean (SD) □ |  | Frame-wise displacement after scrubbing, Mean (SD) □ |  | IQ, Mean (SD) □ |  | Average years between Visits (SD) |
| --- | --- | --- | --- | --- | --- | --- | --- | --- | --- |
| HC | 171 | 120(64/56) | 16.77 (5.80) | p-value: 0.657 | 0.14 (0.06) | p-value <0.001* | 110.69(12.9) | p-value <0.001* | 1.44(2.14) |
| 22q11DS | 160 | 105(50/55) | 17.63 (6.06) |  | 0.18 (0.06) |  | 71.96(13.31) |  | 1.59(2.19) |
| PS+ | 101 | 64(33/31) | 18.15(5.50) | p-value: 0.945 | 0.17(0.06) | p-value: 0.657 | 72.20(14.1) | p-value: 0.729 | 1.63(2.18) |
| PS- | 53 | 35(18/17) | 18.23(5.56) |  | 0.18(0.05) |  | 71.20(12.6) |  | 1.79(2.29) |
| ITS + | 67 | 37(20/17) | 18.43(4.34) | p-value: 0.727 | 0.15(0.04) | p-value: 0.007 * | 67.02(12.4) | p-value: 0.005* | 2.3(2.17) |
| ITS - | 87 | 62(28/34) | 18.02(6.11) |  | 0.19(0.06) |  | 74.72(13.5) |  | 1.31(2.17) |
| ITS + & PS+ | 55 | 30(15/15) | 18.67(4.46) |  | 0.15 (0.05) |  | 67.25(12.4) |  | 2.40(2.27) |

|  | PS+ | PS– | p-value PS+ vs. PS– ◇ | ITS + | ITS - | p-value ITS + vs. ITS – ◇ | 22q1DS |
| --- | --- | --- | --- | --- | --- | --- | --- |
| Anxiety disorder ¥ | 35(54.67%) | 10(28.57%) | 0.008 * | 20(54.05%) | 25(40.32%) | 0.182 | 45 (42.85%) |
| Attention deficit hyperactivity disorder ¥ | 35(54.67%) | 16 (45.71%) | 0.392 | 22(59.46%) | 29(46.77%) | 0.216 | 53 (50.47%) |
| Mood disorder ¥ | 31 (48.44%) | 11 (31.43%) | 0.089 | 19(51.35%) | 23(37.10%) | 0.164 | 44 (41.90%) |
| Schizophrenia ¥ | 6(9.37%) | 0 | NA | 5(13.51%) | 1(1.61%) | 0.047 * | 6 (5.71.4%) |
| More than one psychiatric comorbidity ¥ | 34(53.12%) | 12(34.28%) | 0.09 | 22(59.46%) | 24(38.71%) | 0.041* | 47(44.76%) |
| Mood stabilizer ¥ | 3(4.69%) | 0 | NA | 3(8.11%) | 0 | NA | 3(2.86%) |
| SSRI ¥ | 13 (20.31%) | 4(11.43%) | 0.227 | 11(29.73%) | 6(9.68%) | 0.017* | 18(17.14%) |
| Neuroleptic ¥ | 9(14.06%) | 1(2.86%) | 0.036* | 8(21.62%) | 2(3.23%) | 0.009 * | 10(9.52%) |
| Psychostimulant ¥ | 19(29.69%) | 10(28.57%) | 0.906 | 13(35.13%) | 16(25.81%) | 0.332 | 29(27.61%) |
| Anxiolytic ¥ | 4(6.25%) | 1(2.86%) | 0.412 | 4(10.81%) | 1(1.61%) | 0.085 | 5(4.76%) |

- \* Significant at the level of $p < 0.05$ . (T-test. Otherwise Chi-square indicated by : ◇) - □ Calculated first level average within subject measurements, and then second level across all subjects. - ¥ Positive if present at any of the repetitive assessments.
IQ measurement: Wechsler Intelligence Scale for Children–III (Wechsler, 1991) for children and the Wechsler Adult Intelligence Scale–III (Wechsler, 1997) for adults. The presence of psychiatric disorders: Clinical interview with the patients using the Diagnostic Interview for Children and Adolescents Revised (DICA-R; Reich, 2000) and the Structured Clinical Interview for DSM-IV Axis I Disorders (SCID-I; First et al., 1996).

### Image Acquisition and Preprocessing

During each visit, participants underwent magnetic resonance imaging (MRI) brain scans. Structural and functional MRI images were obtained at the Centre d’Imagerie BioMédicale in Geneva on a Siemens Trio (n = 321) and a Siemens Prisma Fit (MAGNETOM Trio upgrade) (n = 10) 3T scanner (Siemens Corp., Erlangen, Germany). A T1-weighted sequence with 192 slices with a volumetric resolution of 0.86 × 0.86 × 1.1 mm3 was used for the structural image acquisition. Resting-state functional scans were acquired using a T2-weighted sequence (200 frames, voxel size = 1.84 × 1.84 × 3.2 mm3, repetition time = 2.4 s) for 8 minutes (65) during which, participants were instructed to fixate on the white cross on a black screen, not to fall asleep and let their mind wander. Functional images were preprocessed using SPM12 (Wellcome Trust Centre for Neuroimaging, London, UK; http://www.fil.ion.ucl.ac.uk/spm) and an in-lab pipeline (described(66)) using functions of the DPARSF(67) and IBASPM (68) toolboxes. An explanation of the resting-state functional MRI preprocessing pipeline, a list of criteria for the exclusion of subjects from our initial scan pool as well as the detailed list of sequence parameters are available in the supplementary materials.

### Amygdala Subdivisions and Connectivity

Regions of interest were selected using FSL’s Juelich histological atlas (69) based on the probabilistic maps of cytoarchitectonic boundaries (70). Using the study-specific DARTEL template (71) the probabilistic atlas was spatially transformed into the individual subject space. A bilateral mask for each region (BLA, CMA and SFA) was then created including voxels with probability of belonging to the respective subdivision being higher than 50%. The voxel assignments were mutually exclusive and in cases of overlap, the voxel was assigned to the subdivision with the highest probability. For each subject, 3 region of interest (ROI) functional connectivity maps in the individual subject-space were created by computing the Pearson’s correlation coefficient for each voxel’s time course with the average time course inside the respective subdivision. The respective voxel-wise ROI functional connectivity maps were then warped back into study’s DARTEL space and further into the Montreal Neurological Institute space for group-wise comparison (72).

### Partial Least Squares Correlation with Longitudinal Regressors

To optimally investigate the longitudinal changes of maturation in amygdala subdivision connectivity patterns, we employed the partial least squares correlation (PLS-C) (73) via the MATLAB-based publicly available toolbox (https://github.com/MIPLabCH/myPLS). As described in our previous publication (74), we defined the design variables to take into account the longitudinal aspect of brain maturation. In brief, a set of design variables (Y) that are chosen to encode diagnosis (22q11.2DS vs HC or PS (+) vs PS(-) or ITS(+) vs ITS(-)), longitudinal and cross-sectional age, and their interactions are included in one side. The amygdala subdivision connectivity maps per subject (X) was included on the imaging side. These two matrices were used to compute a cross-covariance matrix (R) as R = Y^T^X. The matrix R was then subjected to the singular value decomposition (R = USV^T^), which leads to the computation of latent components (LCs). By projecting each subject’s brain data (X) into the multivariate brain salience (V), we calculate the brain scores as L_X_=XV. These brain scores express how similar each subject’s brain data is to the detected brain salience. Prior to entering the variables into PLS-C, full-scale IQ, sex and motion (average framewise displacement after scrubbing provided by the power formula (75)) were regressed out at a voxel-wise level. A detailed explanation of the inputs to the PLS -C is provided in the supplementary materials.

To assess the significance of the explained correlation for each LC, permutation testing was conducted. In doing so, a null distribution for the singular values was produced using 1000 permutations. Then, for each significant LC the stability of brain and design saliences were assessed using a bootstrapping procedure with 500 random samples with replacement. Of note, to respect all intrasubject dependencies in the longitudinal sample, the random samples were selected across subjects and not scans. The bootstrap ratio for each voxel was calculated as the mean of distribution of all brain saliences for each voxel divided by their respective standard deviation. This value could be interpreted as a measure of stability of each particular voxel in its contribution to the detected LC. In brain maps visualized in this study, bootstrap ratios with an absolute value higher than 3 are depicted, which corresponds to a 99% confidence interval not crossing zero and therefore a stable contribution to the LC. As well, the design saliences are depicted in a bar plot with an error bar showing their respective bootstrap ratio standard deviation.

## Results

For each amygdala subdivision, a first set of PLS-C analysis was conducted to assess the difference of seed to whole-brain functional connectivity maturation between patients with 22q11.2DS and HCs. Subsequently for each subdivision, two follow-up PLS-C analysis was conducted within patients with 22q11.2DS, assessing the difference of functional connectivity maturation between PS(+) and PS(-) patients, as well as ITS(+) and ITS(-) patients. The following sections describe the results of 3 PLS-C analysis within each amygdala subdivision: BLA, CMA and SFA. This processed is summarized and visualized in supplementary materials (Figure S1).

### BLA Connectivity

Upon evaluating developmental differences of the BLA to whole-brain functional connectivity across patients with 22q11.2DS and healthy controls, the PLS-C analysis resulted in one significant component (Pval = 0.001).

This component, as visualized in Fig1-1, captured a substantial effect for diagnosis, along with a relatively smaller effect of maturation as demonstrated by the small age-related loadings in Figure 1-1.a. Overall, the component showed an abnormal pattern of BLA functional connectivity in patients with 22q11.2DS that presented early on during development and slightly deteriorated with age (as visualized in Fig1-1.b).

**Fig 1.**
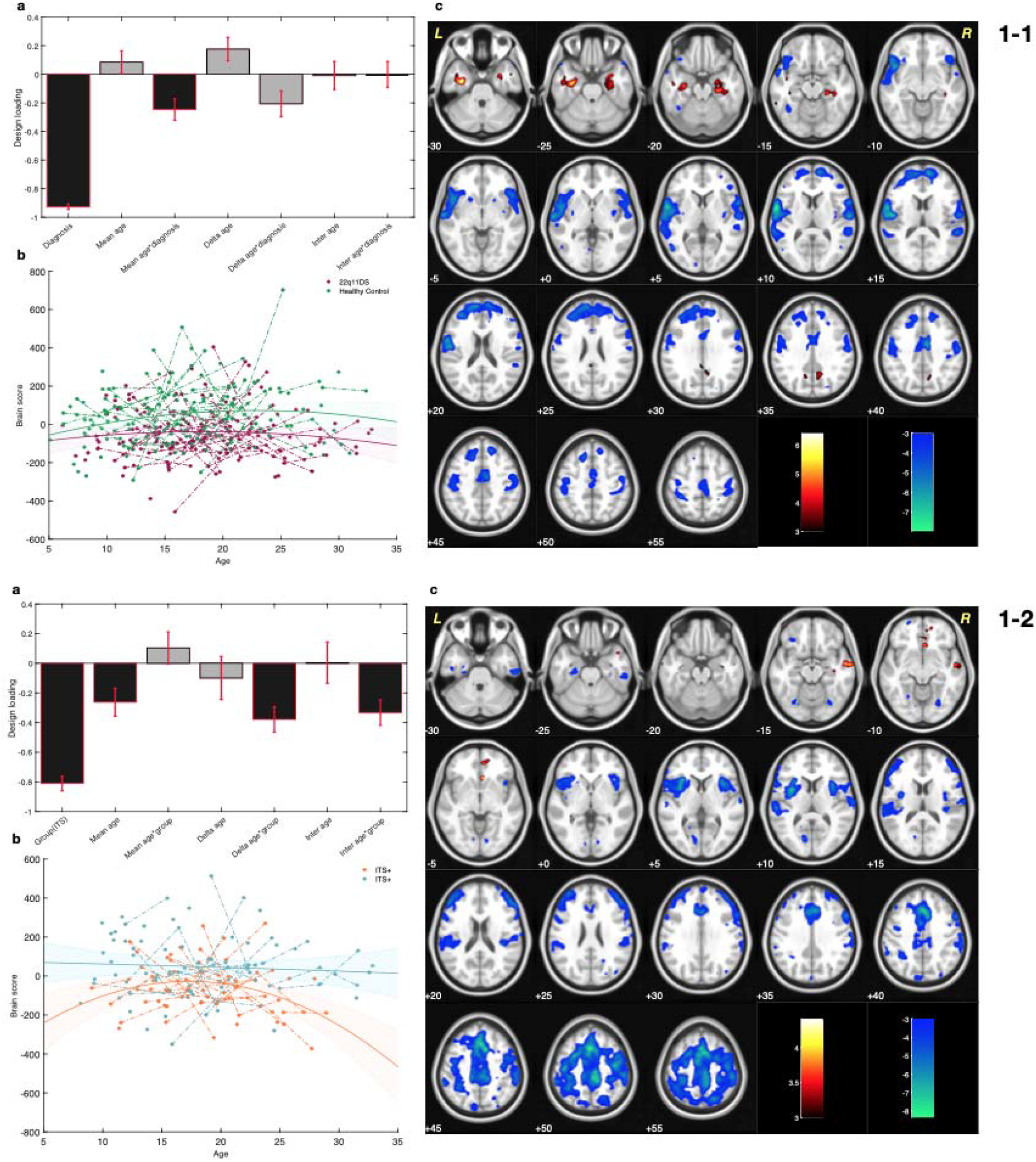
Results of Partial Lease Squares correlation for basolateral amygdala (BLA) functional connectivity at rest. Figure 1-1. Comparing maturation of BLA functional connectivity, in patients with 22q11.2 deletion syndrome (22q11.2DS) and healthy controls subjects. (a) Design salience reveals a substantial negative loading for diagnosis. (b) Distribution of the brain sores across age span for each diagnostic group visualizes the loadings captured in the design salience. Brain scores are lower in patients with 22q11.2DS (negative diagnosis loading). Fitted line and 95% confidence interval are plotted to visualize the age and diagnosis interaction effect captured in the designed salience. (c) The patterns of brain salience could be interpreted as areas with lower BLA functional connectivity (blue), and higher functional connectivity (red) in patients with 22q11.2DS. Figure 1-2 Comparing maturation of BLA functional connectivity in patients with and without mild to moderate impaired tolerance to stress [ITS(+) and ITS(–), respectively]. (a) Design salience reveals the negative effect of group as well as a negative effect of age–group interaction. (b) The distribution of brain scores across age in each group visualizes the captured loadings in the design salience. Accordingly, brain scores are low in ITS(+) patients and deviate further from brain scores of ITS(–) patients during development. Fitted line and 95% confidence interval are plotted to visualize the age and diagnosis interaction effect captured by partial least squares correlation in the design salience. (c) The pattern of brain salience shows areas with lower and more severe decrease of BLA functional connectivity in ITS(+) patients (blue).

The multivariate brain pattern revealed that in patients with 22q11.2DS, the BLA was functionally hyperconnected to hippocampal and para-hippocampal regions (Red clusters in Fig1-1.c). Whereas, in another set of clusters, patients with 22q11.2DS showed abnormally lower functional connectivity with BLA when compared to HC. These hypoconnectivity clusters were located in anterior cingulate cortex, dorsolateral prefrontal cortex (DLPFC), middle prefrontal cortex (mPFC), insula, post-central and superior temporal gyrus (depicted by blue clusters in Fig 1-1.c).

A separate PLS-C analysis was conducted to assess the difference of BLA connectivity among deletion carriers with and without ITS, which resulted in one significant component (Pval = 0.001). As demonstrated in Fig 1-2, this component captured clusters of lowered BLA connectivity within the anterior cingulate cortex, paracingulate gyrus, dorsolateral prefrontal cortex (DLPFC), insula, supplementary motor area (SMA) and post-central gyrus (blue clusters in Fig 1-2.c). As captured by the large loading on ITS in figure 1-2. a, the presence of symptoms of tolerance to stress had a considerable contribution to the component alongside the smaller loadings on age-interaction related loadings. Thus, this component captured a pattern of BLA functional hypoconnectivity that was more severe in patients with ITS and got accentuated as the patients aged (trajectories visualized in Fig 1-2.b).

PLS-C analysis comparing BLA connectivity in 22q11.2DS patients with and without PS resulted in no significant component (LC1, Pval=0.082).

### CMA Connectivity

The PLS-C analysis investigating abnormal development of CMA functional connectivity in patients with 22q11.2DS resulted in one significant component (Pval=0.001). As shown in Fig 2-1.a, this component mainly captured the effect of maturation (stable age-related loadings). As indicated by the considerable loading on age-diagnosis interaction, this component revealed deviations in the developmental trajectory of CMA connectivity in patients with 22q11.2DS when compared to healthy controls (as visualized in Fig2-1.b).

**Fig 2.**
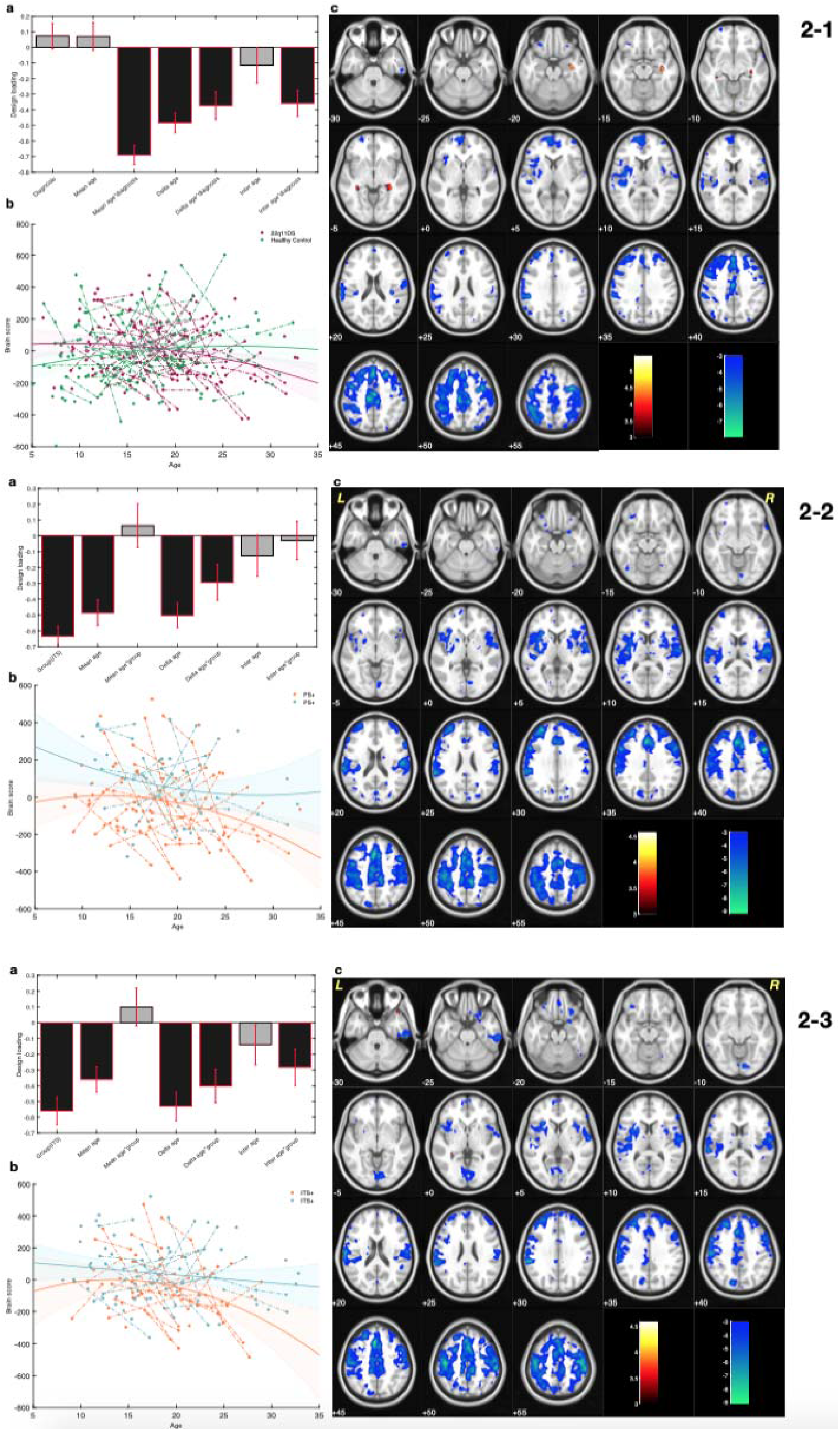
Results of Partial Lease Squares correlation for centromedial amydgala (CMA) functional connectivity at rest. Figure 2-1. Comparing maturation of CMA functional connectivity, in patients with 22q11.2 deletion syndrome (22q11.2DS) and healthy controls subjects. (a) Design salience reveals negative values for age related loadings. (b) Distribution of the brain sores across age for each group visualizes the loadings captured in the design salience. Brain scores progressively decrease with aging in patients with 22q11.2DS but not in healthy controls (negative age-diagnosis interaction loadings). Fitted line and 95% confidence interval are plotted to visualize the age and diagnosis interaction effect captured in the designed salience. (c) The patterns of brain salience could be interpreted as areas that show a progressive decrease in CMA functional connectivity in patients with 22q11.2DS but not healthy controls (blue). Figure 2-2 Comparing maturation of CMA functional connectivity in patients with and without mild to moderate positive psychotic symptoms [PS(+) and PS(–), respectively]. (a) Design salience reveals the negative effect of group as well as s negative effects of age–group interactions. (b) The distribution of brain scores across age in each group visualizes the captured loadings in the design salience. As such, brain scores are low in PS(+) patients and progressively become lower during development when compared to PS(–) patients (negative diagnosis and age-diagnosis interaction loadings). Fitted line and 95% confidence interval are plotted to visualize the loadings captured by partial least squares correlation in the design salience. (c) The pattern of brain salience shows areas with lower and more severe decrease of CMA functional connectivity in PS(+) patients (blue). Figure 2-3 Comparing maturation of CMA functional connectivity in patients with and without mild to moderate impaired tolerance to stress [ITS(+) and ITS(–), respectively]. (a) Design salience reveals the negative effect of ITS group as well as negative effects of age–group interactions. (b) Brain scores distributed across age in each group visualizes the captured loadings in the design salience. Brain scores are low in ITS(+) patients and progressively become lower during development when compared to ITS(–) patients (negative diagnosis and age-diagnosis interaction loadings). Fitted line and 95% confidence interval are plotted to visualize the loadings captured by partial least squares correlation in the design salience. (c) The pattern of brain salience shows a highly comparable brain pattern with Figures 2-1 and 2-2. Likewise this brain pattern visualized areas with lower and more severe decrease of CMA functional connectivity in ITS(+) patients.

The corresponding brain pattern showed clusters within the frontal cortex, insula, SMA and middle cingulate cortex (Blue clusters in Fig2-1.c) that express a developmental drop in functional connectivity with CMA in patients with 22q11.2DS.

Similarly, a second set of PLS-C analysis was conducted to compare differences among patients with and without PS (Fig2-2) resulting in one significant component (Pval=0.002). As revealed by the design loadings (Fig2-2.a), both psychopathology and aging had a considerable effect in this component. Therefore, this component captured decrease in CMA connectivity with clusters in frontal regions, the SMA and the insula (blue clusters in Fig2-2.c). Furthermore, it shows this decrease is most severe in patients with PS and decreases more progressively with aging in this group (as visualized in Fig 2-2.b).

While comparing patients with and without ITS, a highly comparable brain pattern (Fig 2-3) was captured in the significant latent component (Pval = 0.001). The design loading (Fig2-3.a) of this component indicated an effect for age and age-group interaction but not a strong loading for group. The developmental trajectory of these groups is visualized as shown in Fig2-3.b, and shows that patients with symptoms of impaired tolerance undergo a more severe developmental drop in CMA connectivity by adulthood.

### SFA Connectivity

The significant component (Pval=0.003) resulted from a PLS-C analysis comparing SFA functional connectivity in patients with 22q11.2DS and healthy controls is shown in Fig3-1. This component captured an effect for diagnosis and relatively stronger one for aging as indicated by the respective stable loadings (Fig3-1a). The multivariate brain pattern within this component revealed clusters with accelerated hyperconnectivity in patients with 22q11.2DS (Red clusters in Fig3-1.c). Overall, this component captured a developmental increase of SFA connectivity within the caudate, putamen, thalamus and occipital cortex that is slightly increased in patients with 22q11.2DS. This is visualized in trajectories of brain-scores over age in Fig 3-1.b.

**Fig 3.**
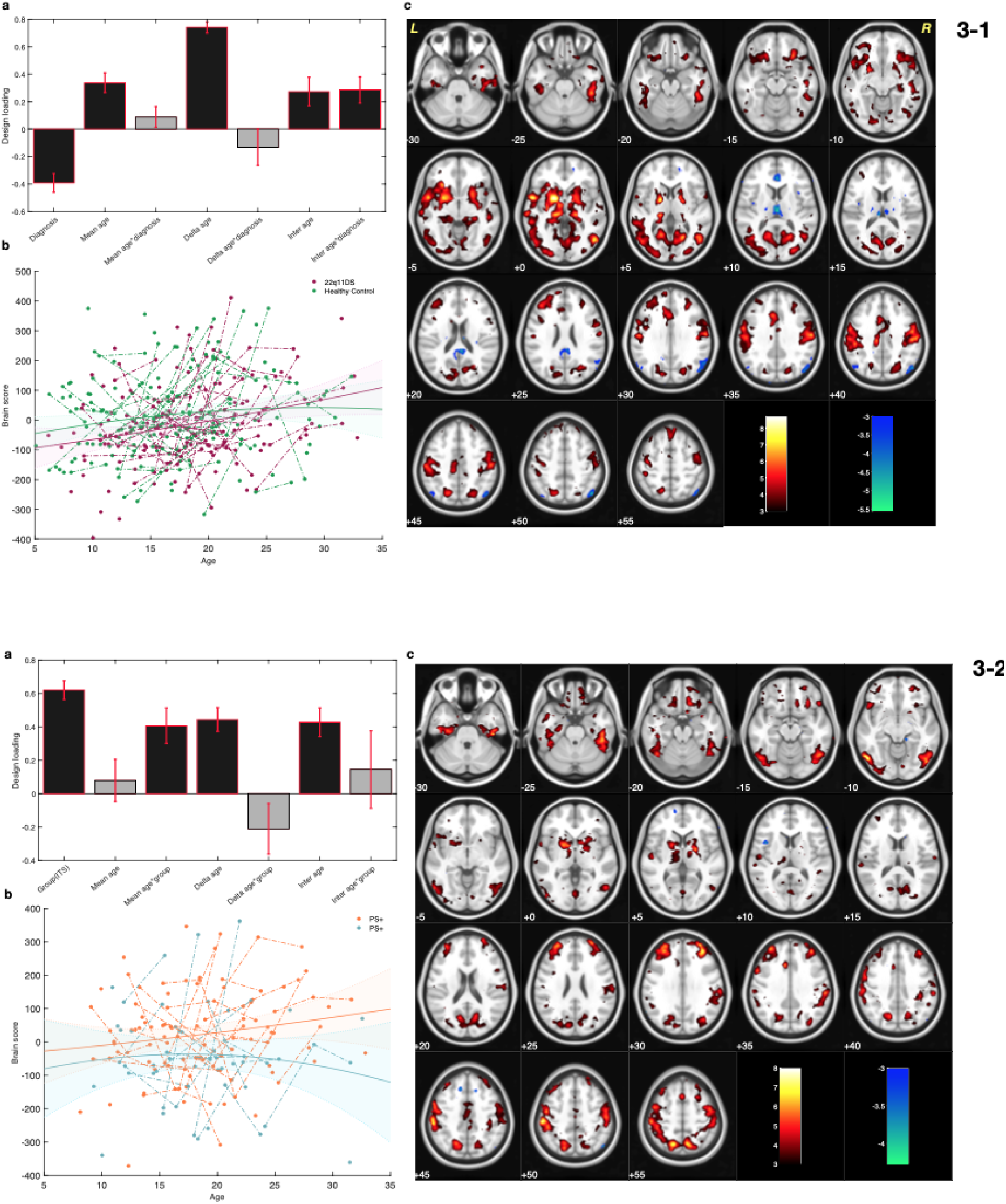
Results of Partial Lease Squares correlation for superficial amygdala (SFA) functional connectivity at rest. Figure 3-1. Comparing maturation of SFA functional connectivity, in patients with 22q11.2 deletion syndrome (22q11.2DS) and healthy controls subjects. (a) Design salience reveals a substantial age-related loading in both groups. (b) Distribution of the brain sores across age span for each diagnostic group visualizes the effect of aging captured in the design salience. Brain scores increase with age (positive age-related loadings). Fitted line and 95% confidence interval are plotted to visualize the loadings captured in the designed salience. (c) The patterns of brain salience could be interpreted as areas with increasing SFA functional connectivity (red), and decreasing SFA functional connectivity (blue). Figure 3-2 Comparing maturation of SFA functional connectivity in patients with and without mild to moderate positive psychotic symptoms [PS(+) and PS(–), respectively]. (a) Design salience reveals the positive effect of group as well as a positive effects for age and age–group interactions. (b) The distribution of brain scores across age in each group visualizes the captured loadings in the design salience. As visualized, brain scores are higher in PS(+) patients and increase faster with aging when compared to the brain scores of PS(–) patients. Fitted line and 95% confidence interval are plotted to visualize the age and diagnosis interaction effect captured by partial least squares correlation in the design salience. (c) The pattern of brain salience shows areas with higher and accelerated increase of SFA functional connectivity in PS(+) patients.

A follow-up set of PLS-C analysis, comparing SFA connectivity in patients with and without PS produced one significant component (Pval =0.010). This component also indicated clusters of hyperconnectivity in the putamen and thalamus (shown in red in fig 3-2.c).

Moreover, the design loading (Fig3-2.a) captured mixed effects of group and age, indicating that, in deletion carriers with PS, the SFA forms stronger functional connections with the striatum and thalamus when compared to deletion carriers without PS.

The PLS-C analysis comparing SFA connectivity in patients with and without ITS produced no significant components.

## Discussion

In the present study we examined the abnormal maturation of resting state connectivity in three subdivisions of amygdala in patients with 22q11.2DS. Using a multivariate longitudinal analysis, we provide the first evidence to date for dysmaturation of amygdala to whole-brain functional connectivity in this population. We further investigated the contribution of amygdala dysconnectivity to positive psychotic symptoms as well as to impaired tolerance to stress in patients carrying the deletion. Figure 4. summarizes our main findings as well as possible underlying mechanisms in which dysfunction of amygdalar subdivision drives stress and psychosis symptoms.

**Figure 4.**
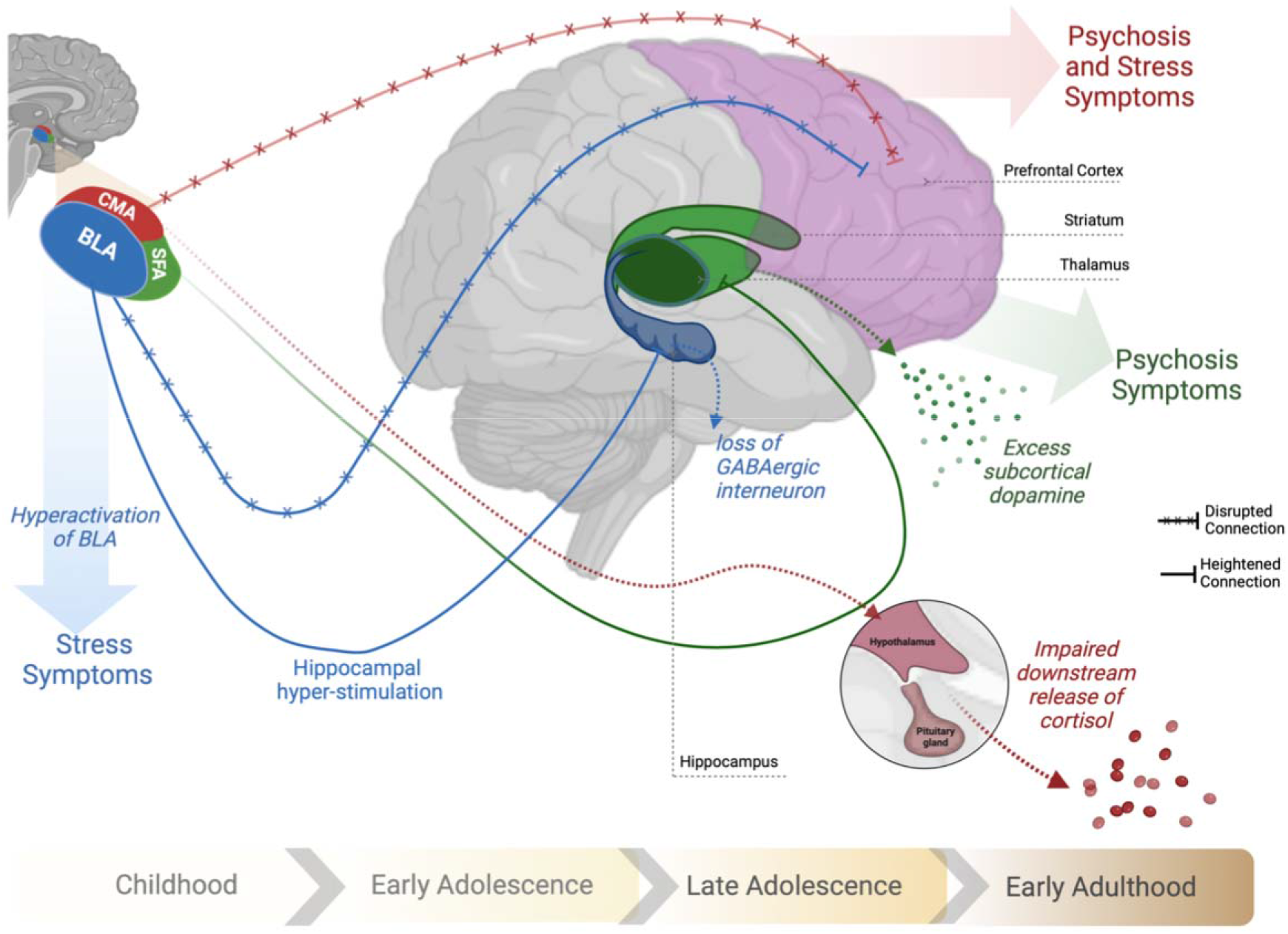
Amygdalar subdivisions and the putative circuits involved in stress and psychosis symptoms in patients with 22q11.2 deletion syndrome. Multivariate pattern of BLA dysconnectivity: During childhood basolateral amygdala (BLA) exhibits impaired connections with prefrontal cortex which would result in hyperactivation of BLA and stress symptoms. Abnormally heightened connectivity to hippocampus modulates loss of GABAergic interneurons in hippocampus that later results in hyperactive hippocampus and positive psychotic symptoms. Centromedial amygdala (CMA) connectivity dysmaturation: a mutual finding in both stress and psychosis symptoms during adolescence. Impaired connections of CMA-prefrontal cortex results in CMA hyperactivation and out of context downstream release of cortisol. Superficial amygdala (SFA) heightened connections to striatum and thalamus: a specific pattern presented with psychotic symptoms possibly through the subcortical excess of dopamine.

The first set of multivariate analysis revealed a pattern of decreased BLA functional connectivity to prefrontal regions as well as the insula and sensory-motor regions. Functionally, BLA is mainly known as a processing hub for the amygdaloid complex by receiving various sensory and associative information from the brainstem, thalamus and the cortex and integrating this information to allow for a proper amygdala response (35, 47).

Specifically, mPFC projections to the BLA form top-down control on the amygdala’s fear and anxiety response (76-78). In fact, the heightened anxiety observed during adolescence in normal development has been attributed to immature connections of mPFC to the BLA (47, 79). By adulthood, the strengthening of BLA to PFC connectivity results in more efficient emotional regulation(76). Consistent with this notion, our analysis showed BLA - PFC connectivity in healthy control increases throughout development (as indicated by positive age-related loadings). However, we found that patients with 22q11.2DS fail to reach this mature state of connectivity. Instead, patients with the deletion showed higher functional connectivity from the BLA to hippocampus and parahippocampus. Dense reciprocal connections between BLA and hippocampus have been previously identified. Notably, impairment of the interconnected circuit between mPFC, hippocampus and BLA had been implicated in anxiety-like behavior (80-83).

Our multivariate results point to higher BLA-hippocampus functional connectivity that co-occurs with lower BLA-PFC functional connectivity in patients with 22q11.2DS. The BLA – hippocampus connections are mainly excitatory(80), whereas BLA inputs to the PFC are predominantly inhibitory(84). Thus, our results may be indicative of BLA hyperactivation in patients with 22q11.2DS, which according to developmental trajectories is already present during childhood. Moreover, BLA-PFC connectivity was particularly lower in patients with moderate to severe ITS. Thus, early dysfunction of the BLA in 22q11.2DS may explain the predisposition to anxiety disorders in childhood in this population. Interestingly, translational studies have shown that BLA activation, whether caused by stress or direct stimulation, disrupts GABAergic inhibitory signaling in the ventral hippocampus (85, 86). Indeed, hippocampal hyperactivation driven by GABAergic dysfunction has been a key finding in schizophrenia and is known to play a major role in psychosis (87, 88). In 22q11.2DS, there is growing evidence of hippocampal disruption contributing to the emergence of positive psychotic symptoms which occurs during adolescence (74, 89, 90). Notably, hippocampal-PFC connections mature during late adolescence whereas BLA-PFC connections mature during pre-adolescence (91, 92). Thus, knowing that BLA hyperactivity modulates hippocampal activity, early dysfunction of the BLA in 22q11.2DS may explain how presence of anxiety disorders in childhood serve as risk factors for the emergence of psychosis later during the development. These results are particularly in line with recent translational evidence showing the role of BLA dysfunction caused by prepubertal stress in causing psychosis-like phenotypes in post-pubertal rats (53). Future longitudinal studies are needed to investigate whether BLA dysconnectivity in childhood can cause hippocampal dysfunction at follow-up in patients with 22q11.2DS.

The multivariate analysis for CMA identified a set of regions with developmental decreases in functional connectivity. The clusters located within the prefrontal cortex and middle cingulate cortex showed an accelerated decrease of CMA functional connectivity in patients with 22q11.2DS when compared to healthy controls. CMA consists of central and medial amygdala nuclei (36, 70), which form the main output of the amygdala to the brainstem and hypothalamus including outputs to the HPA-axis (93). Additionally, CMA connections to the cortex are critical for attributing cortical attention and forming cortical readiness for appropriate brain-stem responses (94). For instance, mPFC connections to CMA have been shown to reduce CMA responsiveness due to forming an inhibitory feedforward circuit (95). In normal development, CMA-frontal connectivity increases from childhood to adulthood (41, 43). Here, we show in patients with 22q11.2DS, values of CMA-prefrontal connectivity diminished during adolescence resulting in lower CMA-prefrontal connectivity by adulthood in patients with 22q11.2DS. Given established interconnections between the CMA and hypothalamic–pituitary–adrenal (HPA) axis, CMA dysconnectivity may be linked to disrupted cortisol response previously documented in this population(96). Additionally, we demonstrate the developmental drop in CMA – frontal connectivity is most severe in patients at a higher risk for positive psychotic symptoms. The accelerated decrease in CMA to frontal connectivity has been previously documented in youth with psychosis (50)and in patients with attenuated psychotic symptoms (51). The loss of connection in this circuit could contribute to the prodromal loss of emotional regulation that often precedes psychosis in schizophrenia(97).

Moreover, when assessing the difference between patients with and without impaired tolerance to stress (ITS), a similar pattern of CMA to frontal dysconnectivity was found. As the CMA is well connected to hypothalamus and brainstem, the CMA-centered hypothesis fits particularly well with prior evidence showing dysregulation of the HPA-axis in patients with 22q11.2DS and its contribution to ITS and overall psychopathology(98). Notably, both our findings regarding CMA-frontal dysconnectivity and prior findings on pituitary dysmaturation seem to be developmentally arising during adolescence. It is worth noting that, impaired tolerance to stress has been indicated as a key symptom in the affective pathway to psychosis in 22q11.2DS (58). Future studies should aim to shed light on how the CMA dysconnectivity and HPA axis dysregulation could interact to contribute to impaired tolerance to stress and psychosis.

Furthermore, our multivariate results point to concurrent hypoconnectivity of the CMA with the somatosensory cortex and insula in PS(+) and ITS(+). Connections of the amygdala to the somatosensory cortex and the insula have been implicated in attributing emotions to sensory stimuli(99, 100). Impairments in somatosensory-centered circuits have also been implicated in emotional embodiment and aberrant environmental appraisal(101). Consequently, sensorimotor dysfunction likely contributes to both affective and psychotic psychopathology (102). Our results regarding CMA to somatosensory dysconnectivity in patients with PS (+) and ITS(+) is in line with evidence showing heightened affective appraisal of sensory stimuli in psychosis (103) and suggests that sensory experiences manifest along the pathway between affective and psychotic symptoms. Overall, our results regarding contribution of CMA dysconnectivity to both PS and ITS, hint toward a key role of the CMA as a neurobiological substrate of affective pathway to psychosis.

Finally, multivariate analysis on SFA-centered connectivity resulted in a pattern of abnormal connectivity specific to psychosis. This included clusters indicating developmental increases in SFA connectivity with the striatum and thalamus that was accelerated in patients with 22q11.2DS with prodromal psychotic symptoms. It must be noted that while developmental increases in amygdala – striatum connectivity in youth with psychosis has been previously reported, it was in regards to the CMA (50). This, aside from differences in the studied population, may be rooted in the use of two amygdala subdivisions (CMA and BLA) in the aforementioned study. Indeed, out-of-context excesses of subcortical dopamine have been one of the major neurochemical hypotheses of psychosis(104). The SFA subdivision in particular includes the anterior amygdaloid area (AAA) which has an embryological striatal origin and forms dense connections to striatum (36, 70). Moreover, upon masking CMA and BLA age-dependent striatum coupling has been found uniquely in SFA (41). On the other hand, the role of the amygdala-striatal pathway has been implicated in transferring aberrant signals to dopaminergic areas (105, 106). Therefore, heightened connections of the SFA to the striatum and thalamus may take part in dopamine dysregulation observed in patients with psychosis.

### Conclusion, Methodological Considerations, and Future Perspectives

The interpretation of our results is restricted by a number of methodological considerations. First, multivariate longitudinal analysis such as the one proposed here allows for data-driven estimation of brain patterns that maximally explain maturation in different groups. While this is a powerful tool in detecting ROI-centered networks undergoing abnormal development, the patterns must be considered as a whole and no conclusions can be drawn for individual connections even if their saliencies are large. Second, the studied population is a mixture of longitudinal and cross-sectional samples–a detailed description of the number of visits in each group is provided in the supplementary material. Indeed, acquiring high-quality longitudinal data of patients with a rare deletion is extremely challenging and this cohort is unique in this regard. In this respect, the flexibility of the longitudinal PLS-C to adapt for each subject to the number of visits makes it particularly well-equipped to take into account the combination of longitudinal and cross-sectional data.

Given the purpose of this study to characterize contributions of the dysmaturation of amygdala subdivisions in different psychopathologies, we did not directly study the effect of sex. Instead, we controlled for the effect of sex by regressing sex out from the connectivity matrix. Undoubtedly, sex differences in the amygdala could provide insightful information and future studies with bigger sample size in both sex groups are needed to investigate how sex may affect the developmental trajectories of amygdala subdivisions connectivity.

Finally, the rate of several psychopathologies and medication intake was higher in patients with PS(+) and ITS(+). Our post-hoc investigation did not reveal any significant association between said variables and brain-scores, as described in detail in Supplementary material. This suggests that our results were not directly driven by the difference observed in these variables. However, more extensive studies are needed to investigate the effect of different classes of medication in amygdala-centered circuits.

In brief, we examined the developmental functional-connectivity patterns of three amygdala subdivisions in participants with 22q11.2DS as well as their respective contributions to positive psychotic symptoms and impaired tolerance to stress. Indeed, each amygdala subdivision showed distinct patterns of dysconnectivity with unique contributions to symptoms of psychopathology we investigated. This further confirms that amygdala subdivisions should be considered separately in terms of functional connectivity if the goal is to disentangle complex psychopathological pathways.

We propose early BLA dysfunction as an early signature in 22q11.2DS which contributes to impaired tolerance to daily stress. In light of previous translational findings, future studies should investigate whether BLA dysfunction in childhood could result in hippocampal dysfunction, and consecutively psychosis, during adolescence. Moreover, we aimed to delineate if abnormal development of amygdala connectivity holds a direct link to psychosis, or whether it contributes to the “affective pathway” to psychosis by symptoms of emotional dysregulation. Our results rather point to a combination of both. Namely, SFA hyperconnectivity to the striatum was revealed as a specific pattern arising in patients who develop mild to moderate positive psychotic symptoms. Conversely, CMA showed frontal dysconnectivity as a mutual neurobiological substrate in both impaired tolerance to stress and psychosis indicating a role in prodromal dysregulation of emotions in psychosis. Further studies should explore how CMA dysconnectivity could interact with HPA axis in impairment of affective pathways and their contributions to conversion to psychosis.

## Data Availability

All data produced in the present study are available upon reasonable request to the authors

## Acknowledgments

This has been supported by the Swiss National Science Foundation (Grant Nos. 324730_121996 and 324730_144260 [to SE]) and a National Centre of Competence in Research Synapsy grant (Grant No. 51NF40-158776 [to SE]).

We express our outmost gratitude to all the families who participated in our study. A special thanks to Tereza Kotalova for coordinating the project and to the MRI operators at the Campus Biotech, Roberto Martuzzi and Loan Mattera. Finally, we would like to thank Karin Bortolin, Valentina Mancini, Niveettha Thillainathan, and Léa Céline Marécaille for their kind help during data collection. Figure 4 is created with BioRender.com.

The authors report no conflicts of interest.

