## Supplemental Material for "Amygdala subdivisions exhibit aberrant whole-brain functional connectivity in relation to stress intolerance and psychotic symptoms in 22q11.2DS"

**Supplemental Methods and Materials**

**Participants**

Over all 451, resting-state fMRI scans were obtained during the ongoing Geneva cohort of 22q11DS. From this initial sample pool, 120 needed to be excluded following scan quality control. This in detail includes: 44 scans which were removed due to excessive movement (over 3mm translation or 3 degrees of rotation), 6 scans that had to be removed because the subject had fallen asleep during the scan and 36 scans that were not used since part of the brain (vertex or the temporal lobe) was cut from the scanning area. Of the remaining scans, after motion scrubbing, 34 had less than 100 rs-fMRI scans with frame-wise displacement of below 0.5 and did not meet the cut-off of 4-minutes to be included in the study. No scan had to be excluded upon visual inspection of the seed mask placement. All exclusions took place prior to analysis of region of interest and connectivity matrices eliminating chances of biased exclusion.

**Table S1: Subject scan-visit per group**

| Number of repetitive scans | HC | 22q11DS | PS+ | PS - | ITS + | ITS - | ITS+ and PS + |
| --- | --- | --- | --- | --- | --- | --- | --- |
| 1 | 78 | 64 | 38 | 20 | 15 | 43 | 12 |
| 2 | 33 | 29 | 17 | 12 | 15 | 14 | 12 |
| 3 | 9 | 10 | 7 | 3 | 6 | 4 | 5 |
| 4 | 0 | 2 | 2 | 0 | 1 | 1 | 1 |

**MRI sequence parameters:**

T2 weighted (200 frames, acquisition matrix = 94 × 128, field of view = 96 × 128, voxel size = 1.84 × 1.84 × 3.2 mm3, 38 axial slices, slice thickness = 3.2 mm, repetition time = 2400 ms, echo time = 30 ms, flip angle = 85°, phase encoding anterior > posterior, descending sequential ordering, GRAPPA [generalized autocalibrating partially parallel acquisitions] acceleration mode with factor for parallel imaging = 2).

T1 weighted (repetition time = 2500 ms, echo time = 3 ms, field of view = 23.5 cm2, flip angle = 8°, acquisition matrix = 256 × 256, slice thickness = 1.1 mm, phase encoding right > left, no fat suppression).

**Preprocessing pipeline description**

Following the realignment of the functional scans we applied spatial smoothing with full width half maximum (FWHM) = 6 mm. We then Co-registered the mean of structural scans and the mean of functional scans. The segmentation of the structural images was performed with the SPM12 Segmentation algorithm (Ashburner and Friston, 2005). To create a template space specific to our population, Diffeomorphic Anatomical Registration using Exponential Lie algebra (DARTEL; Ashburner, 2007) was used to compute the study- specific template. Voxel-wise time-series were extracted in the native space and first five functional scans were excluded from the analysis. The BOLD time series were then regressed out for mean white-matter and CSF signals. The band-pass filtering of 0.01 Hz to 0.1 Hz was then applied. Motion scrubbing (Power et al., 2012), was then applied with a frame-wise displacement of more than 0.5mm for the selected frame as well as a frame before and two frames after.

**A detailed explanation of the inputs to the PLS -C**

For the design variables included into the PLS-C, we start by including a binary variable of diagnosis, which is (depending on the analysis) either HCs versus patients with 22q11DS, PS(+) versus PS(−) or ITS(+) versus ITS(-). Given the longitudinal design of the study, we designed the age-related variables as described below to be able to capture the effect of aging both over the whole cohort as well as the inter-subject effect of age.

First, mean-age which is the average age of that subject across all visits included in the study. Next, delta-age assigns the difference between the actual age of a subject at a visit and that subject’s mean-age. As described in our previous publication, by construction, the orthogonalized mean-age and delta-age correspond to cross-sectional and longitudinal effect of age, respectively. Last, inter-age is defined as the interaction between mean-age and delta-age, to characterize the existence of a U-shaped curve in the neurodevelopmental trajectories.

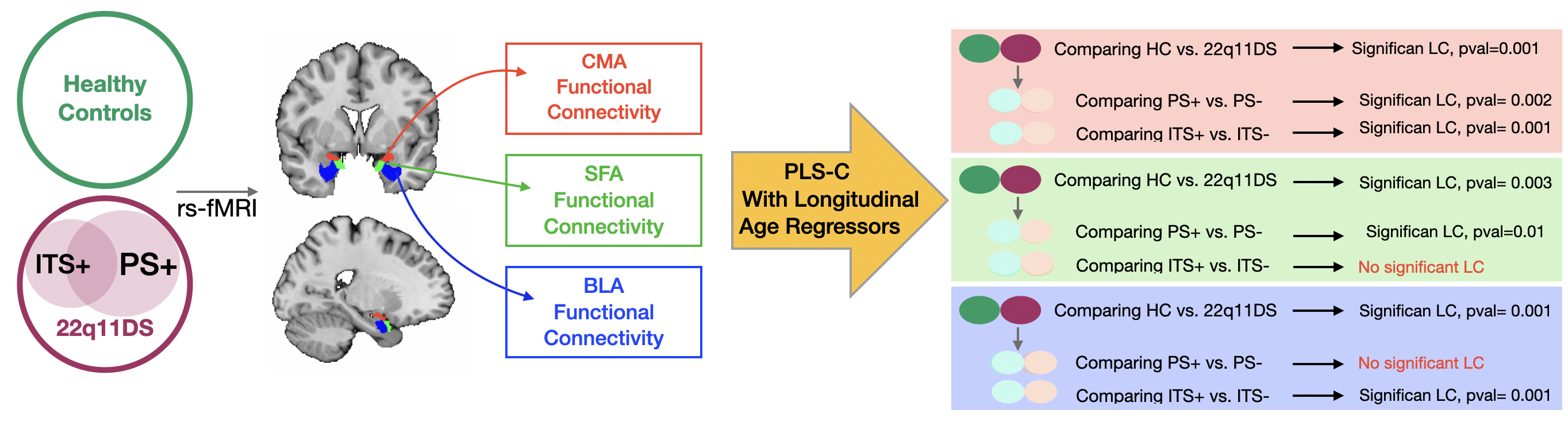

*Figure S1. an overview of the analysis pipeline and results*

**Supplemental Results**

**Post-hoc analysis on possible confounding factors**

According to Table. 1 several rates of psychopathology and intake of medication were different among PS and ITS groups. A follow-up post-hoc analysis using partial-correlation was carried out to investigate the contribution of each of these differences in the brain-scores obtained from PLS-C in a cross-sectional sample.

**Table S2: Partial correlation of brain-score and intake of medication and diagnosis of psychopathology**

| Brain-score of cross-sectional samples (n=99) | | Anxiety disorder, Rho (p-val) | Schizophrenia, Rho (p-val) | SSRI, Rho (p-val) | Neuroleptic, Rho (p-val) |
| --- | --- | --- | --- | --- | --- |
| BLA | ITS+ vs ITS- | 0.070(0.49) | -0.057(0.58) | -0.139(0.17) | -0.023(0.82) |
| CMA | PS+ vs PS- | 0.100(0.33) | 0.035(0.73) | -0.158(0.12) | -0.026(0.80) |
|  | ITS+ vs ITS- | 0.073(0.48) | 0.035(0.73) | -0.107(0.30) | -0.064(0.53) |
| SFA | PS+ vs PS- | 0.140(0.17) | -0.044(0.67) | -0.050(0.63) | 0.096(0.35) |

**Brain Maps Bootstrap Ratio Per Area:**

| **BLA 22q11DS vs HC** | | | | |
| --- | --- | --- | --- | --- |
| **ROI size (Voxel)** | **Cluster size (Voxels)** | **Max**  **(+/-) value** | **Coverage** | **Region Label** |
| 978 | 395 | -7.8260984 | 0.40388548 | CO l (Central Opercular Cortex Left) |
| 698 | 211 | 5.8691802 | 0.30229226 | Hippocampus r |
| 319 | 81 | 5.98949909 | 0.2539185 | pPaHC r (Parahippocampal Gyrus, posterior division Right) |
| 392 | 93 | -6.3761425 | 0.2372449 | pSTG l (Superior Temporal Gyrus, posterior division Left) |
| 280 | 62 | -6.3784881 | 0.22142857 | aSTG l (Superior Temporal Gyrus, anterior division Left) |
| 761 | 144 | 6.30982113 | 0.1892247 | Hippocampus l |
| 342 | 63 | 5.70457363 | 0.18421053 | Amygdala r |
| 656 | 103 | 5.19016743 | 0.1570122 | aPaHC r (Parahippocampal Gyrus, anterior division Right) |
| 877 | 108 | -6.6559577 | 0.12314709 | CO r (Central Opercular Cortex Right) |
| 565 | 68 | -6.5895686 | 0.12035398 | PT l (Planum Temporale Left) |
| 440 | 38 | -6.3430929 | 0.08636364 | PT r (Planum Temporale Right) |
| 860 | 72 | 6.2514596 | 0.08372093 | pTFusC l (Temporal Fusiform Cortex, posterior division Left) |
| 766 | 64 | -6.1324382 | 0.08355091 | IFG oper l (Inferior Frontal Gyrus, pars opercularis Left) |
| 355 | 28 | -6.6004796 | 0.07887324 | FO l (Frontal Operculum Cortex Left) |
| 317 | 25 | 7.16636372 | 0.07886435 | aTFusC l (Temporal Fusiform Cortex, anterior division Left) |
| 4359 | 335 | -8.6905184 | 0.07685249 | PreCG l (Precentral Gyrus Left) |
| 719 | 55 | 5.52675486 | 0.07649513 | pTFusC r (Temporal Fusiform Cortex, posterior division Right) |
| 2593 | 193 | -7.2247357 | 0.07443116 | AC (Cingulate Gyrus, anterior division) |
| 1680 | 121 | -6.9540162 | 0.07202381 | FOrb l (Frontal Orbital Cortex Left) |
| 327 | 23 | 5.56582212 | 0.07033639 | Amygdala l |
| 292 | 20 | 4.80510807 | 0.06849315 | aTFusC r (Temporal Fusiform Cortex, anterior division Right) |
| 6926 | 457 | -6.607759 | 0.06598325 | FP l (Frontal Pole Left) |
| 309 | 18 | -5.9513764 | 0.05825243 | HG l (Heschl's Gyrus Left) |
| 582 | 33 | 6.66414309 | 0.05670103 | aPaHC l (Parahippocampal Gyrus, anterior division Left) |
| 409 | 19 | 3.88363719 | 0.04645477 | aMTG r (Middle Temporal Gyrus, anterior division Right) |
| 1235 | 51 | -5.7881694 | 0.04129555 | pSMG r (Supramarginal Gyrus, posterior division Right) |
| 714 | 28 | -6.5407181 | 0.03921569 | SMA r (Juxtapositional Lobule Cortex -formerly Supplementary Motor Cortex- Right) |
| 686 | 26 | -6.1364617 | 0.03790087 | IFG oper r (Inferior Frontal Gyrus, pars opercularis Right) |
| 1442 | 53 | -5.8397245 | 0.03675451 | FOrb r (Frontal Orbital Cortex Right) |
| 650 | 21 | -5.9685183 | 0.03230769 | IFG tri l (Inferior Frontal Gyrus, pars triangularis Left) |
| 313 | 10 | -6.0715642 | 0.03194888 | FO r (Frontal Operculum Cortex Right) |
| 3657 | 108 | -8.800189 | 0.0295324 | PostCG l (Postcentral Gyrus Left) |
| 5613 | 146 | 4.75489902 | 0.02601105 | Precuneous (Precuneous Cortex) |
| 555 | 14 | -5.7482018 | 0.02522523 | IFG tri r (Inferior Frontal Gyrus, pars triangularis Right) |
| 2042 | 41 | -5.9700084 | 0.02007835 | iLOC l (Lateral Occipital Cortex, inferior division Left) |
| 4275 | 83 | -7.9149804 | 0.0194152 | PreCG r (Precentral Gyrus Right) |
| 2831 | 51 | -5.6375165 | 0.01801484 | SFG l (Superior Frontal Gyrus Left) |
| 8081 | 135 | -4.5008688 | 0.01670585 | FP r (Frontal Pole Right) |
| 3233 | 49 | -7.8753705 | 0.0151562 | PostCG r (Postcentral Gyrus Right) |
| 336 | 5 | 4.09126091 | 0.01488095 | aITG l (Inferior Temporal Gyrus, anterior division Left) |
| 698 | 10 | 3.39139652 | 0.01432665 | toITG l (Inferior Temporal Gyrus, temporooccipital part Left) |
| 1020 | 14 | 3.73320222 | 0.01372549 | pITG l (Inferior Temporal Gyrus, posterior division Left) |
| 1270 | 17 | 3.95081782 | 0.01338583 | Thalamus r |
| 390 | 5 | 3.74809957 | 0.01282051 | pPaHC l (Parahippocampal Gyrus, posterior division Left) |
| 643 | 7 | -4.7814722 | 0.01088647 | SMA L(Juxtapositional Lobule Cortex -formerly Supplementary Motor Cortex- Left) |

| **CMA 22q11DS vs HC** | | | | |
| --- | --- | --- | --- | --- |
| **ROI size (Voxel)** | **Cluster size (Voxels)** | **Max**  **(+/-) Value** | **Coverage** | **Region Label** |
| 954 | 567 | -7.1248636 | 0.59433962 | aSMG l (Supramarginal Gyrus, anterior division Left) |
| 1460 | 861 | -7.4211307 | 0.58972603 | SPL l (Superior Parietal Lobule Left) |
| 643 | 379 | -7.2915101 | 0.58942457 | SMA L(Juxtapositional Lobule Cortex -formerly Supplementary Motor Cortex- Left) |
| 309 | 127 | -6.0204449 | 0.41100324 | HG l (Heschl's Gyrus Left) |
| 2927 | 1163 | -7.1607366 | 0.39733516 | MidFG l (Middle Frontal Gyrus Left) |
| 567 | 194 | -6.7289324 | 0.34215168 | PO l (Parietal Operculum Cortex Left) |
| 2831 | 933 | -7.8753638 | 0.32956552 | SFG l (Superior Frontal Gyrus Left) |
| 1476 | 485 | -7.2816448 | 0.32859079 | SPL r (Superior Parietal Lobule Right) |
| 3657 | 1097 | -7.745286 | 0.29997266 | PostCG l (Postcentral Gyrus Left) |
| 1313 | 388 | -8.6204195 | 0.29550647 | PaCiG l (Paracingulate Gyrus Left) |
| 3233 | 935 | -7.0849042 | 0.28920507 | PostCG r (Postcentral Gyrus Right) |
| 4359 | 1235 | -9.6522179 | 0.28332186 | PreCG l (Precentral Gyrus Left) |
| 978 | 273 | -6.4704342 | 0.2791411 | CO l (Central Opercular Cortex Left) |
| 714 | 197 | -7.6060743 | 0.27591036 | SMA r (Juxtapositional Lobule Cortex -formerly Supplementary Motor Cortex- Right) |
| 2740 | 718 | -7.9750872 | 0.2620438 | MidFG r (Middle Frontal Gyrus Right) |
| 4275 | 1024 | -8.8873653 | 0.23953216 | PreCG r (Precentral Gyrus Right) |
| 1334 | 306 | -7.6636186 | 0.22938531 | IC l (Insular Cortex Left) |
| 1361 | 312 | -7.4963431 | 0.2292432 | PaCiG r (Paracingulate Gyrus Right) |
| 565 | 118 | -6.8080482 | 0.20884956 | PT l (Planum Temporale Left) |
| 2397 | 496 | -8.7308111 | 0.20692532 | PC (Cingulate Gyrus, posterior division) |
| 867 | 147 | -6.1030788 | 0.16955017 | Putamen l |
| 801 | 134 | -6.488976 | 0.16729089 | aSMG r (Supramarginal Gyrus, anterior division Right) |
| 1064 | 174 | -6.8637195 | 0.16353383 | pSMG l (Supramarginal Gyrus, posterior division Left) |
| 2680 | 399 | -7.4332519 | 0.1488806 | SFG r (Superior Frontal Gyrus Right) |
| 4967 | 727 | -6.6406422 | 0.14636602 | sLOC l (Lateral Occipital Cortex, superior division Left) |
| 6926 | 985 | -6.7332745 | 0.14221773 | FP l (Frontal Pole Left) |
| 877 | 119 | -5.8151283 | 0.13568985 | CO r (Central Opercular Cortex Right) |
| 355 | 40 | -4.6221905 | 0.11267606 | FO l (Frontal Operculum Cortex Left) |
| 950 | 92 | -5.9849739 | 0.09684211 | AG l (Angular Gyrus Left) |
| 2593 | 249 | -8.4983349 | 0.09602777 | AC (Cingulate Gyrus, anterior division) |
| 538 | 50 | -6.6448936 | 0.0929368 | PO r (Parietal Operculum Cortex Right) |
| 766 | 63 | -5.1378922 | 0.08224543 | IFG oper l (Inferior Frontal Gyrus, pars opercularis Left) |
| 1235 | 88 | -5.8706765 | 0.07125506 | pSMG r (Supramarginal Gyrus, posterior division Right) |
| 4813 | 313 | -6.8063641 | 0.0650322 | sLOC r (Lateral Occipital Cortex, superior division Right) |
| 5613 | 360 | -6.4903383 | 0.06413683 | Precuneous (Precuneous Cortex) |
| 8081 | 442 | -6.1540737 | 0.0546962 | FP r (Frontal Pole Right) |
| 1344 | 52 | -5.3298407 | 0.03869048 | IC r (Insular Cortex Right) |
| 946 | 36 | -5.0345073 | 0.03805497 | pITG r (Inferior Temporal Gyrus, posterior division Right) |
| 698 | 25 | 5.07637215 | 0.03581662 | Hippocampus r |
| 440 | 15 | -4.9422245 | 0.03409091 | PT r (Planum Temporale Right) |
| 1469 | 49 | -6.0194278 | 0.03335602 | AG r (Angular Gyrus Right) |
| 650 | 21 | -4.6873593 | 0.03230769 | IFG tri l (Inferior Frontal Gyrus, pars triangularis Left) |
| 282 | 8 | -4.9004445 | 0.02836879 | HG r (Heschl's Gyrus Right) |
| 686 | 17 | -4.8661575 | 0.02478134 | IFG oper r (Inferior Frontal Gyrus, pars opercularis Right) |
| 536 | 13 | -5.0935135 | 0.02425373 | Caudate l |
| 1680 | 29 | -4.7796097 | 0.0172619 | FOrb l (Frontal Orbital Cortex Left) |
| 1365 | 21 | -5.6960139 | 0.01538462 | Thalamus l |
| 392 | 4 | -4.3095937 | 0.01020408 | pSTG l (Superior Temporal Gyrus, posterior division Left) |

| **SFA 22q11DS vs HC** | | | | |
| --- | --- | --- | --- | --- |
| **ROI size (Voxel)** | **Cluster size (Voxels)** | **Max**  **(+/-)**  **Value** | **Coverage** | **Region Label** |
| 302 | 148 | 10.4093494 | 0.49006623 | Pallidum l |
| 269 | 115 | 9.94943237 | 0.42750929 | Pallidum r |
| 867 | 344 | 9.48209667 | 0.39677047 | Putamen l |
| 804 | 230 | 9.06342793 | 0.28606965 | Putamen r |
| 752 | 201 | 6.60704327 | 0.26728723 | ICC r (Intracalcarine Cortex Right) |
| 946 | 177 | 6.70557833 | 0.18710359 | pITG r (Inferior Temporal Gyrus, posterior division Right) |
| 2680 | 498 | 10.3187056 | 0.1858209 | SFG r (Superior Frontal Gyrus Right) |
| 714 | 127 | 9.64740562 | 0.17787115 | SMA r (Juxtapositional Lobule Cortex -formerly Supplementary Motor Cortex- Right) |
| 2042 | 348 | 6.5526619 | 0.17042116 | iLOC l (Lateral Occipital Cortex, inferior division Left) |
| 2045 | 342 | 7.61695671 | 0.16723716 | iLOC r (Lateral Occipital Cortex, inferior division Right) |
| 4359 | 714 | 7.72156715 | 0.16379904 | PreCG l (Precentral Gyrus Left) |
| 2831 | 439 | 7.8362689 | 0.15506888 | SFG l (Superior Frontal Gyrus Left) |
| 4275 | 642 | 7.29986525 | 0.15017544 | PreCG r (Precentral Gyrus Right) |
| 3233 | 479 | 7.12994099 | 0.1481596 | PostCG r (Postcentral Gyrus Right) |
| 1365 | 185 | 6.61164999 | 0.13553114 | Thalamus l |
| 641 | 83 | 5.73639488 | 0.12948518 | ICC l (Intracalcarine Cortex Left) |
| 643 | 83 | 6.46884155 | 0.12908243 | SMA L(Juxtapositional Lobule Cortex -formerly Supplementary Motor Cortex- Left) |
| 392 | 49 | 6.00231266 | 0.125 | pSTG l (Superior Temporal Gyrus, posterior division Left) |
| 1334 | 166 | 6.54606199 | 0.12443778 | IC l (Insular Cortex Left) |
| 3657 | 446 | 6.98767281 | 0.12195789 | PostCG l (Postcentral Gyrus Left) |
| 719 | 78 | 6.99331474 | 0.10848401 | pTFusC r (Temporal Fusiform Cortex, posterior division Right) |
| 358 | 35 | 6.6690979 | 0.09776536 | PP l (Planum Polare Left) |
| 73 | 7 | 4.85264158 | 0.09589041 | SCC l (Supracalcarine Cortex Left) |
| 954 | 82 | 6.25581837 | 0.08595388 | aSMG l (Supramarginal Gyrus, anterior division Left) |
| 280 | 20 | 5.50236368 | 0.07142857 | aSTG l (Superior Temporal Gyrus, anterior division Left) |
| 860 | 49 | 5.87885952 | 0.05697674 | pTFusC l (Temporal Fusiform Cortex, posterior division Left) |
| 781 | 44 | 5.63060427 | 0.05633803 | toITG r (Inferior Temporal Gyrus, temporooccipital part Right) |
| 866 | 48 | 5.43987751 | 0.05542725 | toMTG l (Middle Temporal Gyrus, temporooccipital part Left) |
| 801 | 42 | 5.86250257 | 0.05243446 | aSMG r (Supramarginal Gyrus, anterior division Right) |
| 536 | 28 | 6.07313776 | 0.05223881 | Caudate l |
| 1680 | 84 | 6.53321171 | 0.05 | FOrb l (Frontal Orbital Cortex Left) |
| 1442 | 68 | 5.96738672 | 0.04715673 | FOrb r (Frontal Orbital Cortex Right) |
| 2593 | 117 | 6.01703691 | 0.04512148 | AC (Cingulate Gyrus, anterior division) |
| 1270 | 54 | 5.74833059 | 0.04251969 | Thalamus r |
| 1732 | 70 | 5.77390432 | 0.0404157 | LG r (Lingual Gyrus Right) |
| 519 | 20 | 5.10827446 | 0.03853565 | Cuneal l (Cuneal Cortex Left) |
| 1313 | 49 | 5.77659035 | 0.03731912 | PaCiG l (Paracingulate Gyrus Left) |
| 978 | 35 | 6.8075242 | 0.03578732 | CO l (Central Opercular Cortex Left) |
| 1516 | 54 | 5.60861874 | 0.03562005 | LG l (Lingual Gyrus Left) |
| 1382 | 44 | 5.97115755 | 0.03183792 | pMTG l (Middle Temporal Gyrus, posterior division Left) |
| 2927 | 93 | 7.16910791 | 0.03177315 | MidFG l (Middle Frontal Gyrus Left) |
| 2366 | 69 | 6.7799325 | 0.02916314 | TP l (Temporal Pole Left) |
| 766 | 21 | 5.70832396 | 0.02741514 | IFG oper l (Inferior Frontal Gyrus, pars opercularis Left) |
| 2397 | 63 | 5.79481602 | 0.02628285 | PC (Cingulate Gyrus, posterior division) |
| 6926 | 158 | 6.4423995 | 0.02281259 | FP l (Frontal Pole Left) |
| 355 | 8 | 5.34823799 | 0.02253521 | FO l (Frontal Operculum Cortex Left) |
| 1357 | 28 | 6.16385031 | 0.02063375 | pMTG r (Middle Temporal Gyrus, posterior division Right) |
| 292 | 6 | 5.19982433 | 0.02054795 | aTFusC r (Temporal Fusiform Cortex, anterior division Right) |
| 1020 | 20 | 5.13232231 | 0.01960784 | pITG l (Inferior Temporal Gyrus, posterior division Left) |
| 2740 | 52 | 7.57444239 | 0.0189781 | MidFG r (Middle Frontal Gyrus Right) |
| 1162 | 20 | 5.68538713 | 0.0172117 | toMTG r (Middle Temporal Gyrus, temporooccipital part Right) |
| 815 | 14 | 5.25011492 | 0.01717791 | TOFusC r (Temporal Occipital Fusiform Cortex Right) |
| 143 | 2 | 5.08680677 | 0.01398601 | SCC r (Supracalcarine Cortex Right) |
| 641 | 8 | 4.81046343 | 0.0124805 | Cuneal r ( Cortex Right) |
| 5613 | 63 | 5.84252548 | 0.01122394 | Precuneous (Precuneous Cortex) |
| 2640 | 27 | 5.39471436 | 0.01022727 | OP l (Occipital Pole Left) |

| **BLA ITS(+) vs ITS(-)** | | | | |
| --- | --- | --- | --- | --- |
| **ROI size (Voxel)** | **Cluster size (Voxels)** | **Max**  **(+/-) value** | **Coverage** | **Region Label** |
| 567 | 340 | -6.6205859 | 0.59964727 | PO l (Parietal Operculum Cortex Left) |
| 643 | 366 | -8.0652189 | 0.56920684 | SMA L(Juxtapositional Lobule Cortex -formerly Supplementary Motor Cortex- Left) |
| 565 | 258 | -7.6433764 | 0.45663717 | PT l (Planum Temporale Left) |
| 801 | 319 | -6.8156853 | 0.39825218 | aSMG r (Supramarginal Gyrus, anterior division Right) |
| 538 | 207 | -7.9676652 | 0.38475836 | PO r (Parietal Operculum Cortex Right) |
| 714 | 224 | -8.2414761 | 0.31372549 | SMA r (Juxtapositional Lobule Cortex -formerly Supplementary Motor Cortex- Right) |
| 1361 | 423 | -8.8472624 | 0.31080088 | PaCiG r (Paracingulate Gyrus Right) |
| 1313 | 324 | -9.0949268 | 0.24676314 | PaCiG l (Paracingulate Gyrus Left) |
| 978 | 209 | -7.6421466 | 0.21370143 | CO l (Central Opercular Cortex Left) |
| 355 | 75 | -6.6887712 | 0.21126761 | FO l (Frontal Operculum Cortex Left) |
| 954 | 191 | -7.4775648 | 0.20020964 | aSMG l (Supramarginal Gyrus, anterior division Left) |
| 1460 | 285 | -6.6274319 | 0.19520548 | SPL l (Superior Parietal Lobule Left) |
| 1334 | 253 | -8.741642 | 0.18965517 | IC l (Insular Cortex Left) |
| 3233 | 580 | -7.853488 | 0.17939994 | PostCG r (Postcentral Gyrus Right) |
| 4275 | 762 | -8.1292982 | 0.17824561 | PreCG r (Precentral Gyrus Right) |
| 313 | 55 | -6.637217 | 0.17571885 | FO r (Frontal Operculum Cortex Right) |
| 278 | 40 | 4.456439972 | 0.14388489 | aSTG r (Superior Temporal Gyrus, anterior division Right) |
| 1476 | 207 | -6.598012 | 0.1402439 | SPL r (Superior Parietal Lobule Right) |
| 4359 | 548 | -7.3039594 | 0.12571691 | PreCG l (Precentral Gyrus Left) |
| 888 | 110 | -6.7605996 | 0.12387387 | OFusG r (Occipital Fusiform Gyrus Right) |
| 2680 | 321 | -7.8021417 | 0.11977612 | SFG r (Superior Frontal Gyrus Right) |
| 440 | 49 | -7.6344724 | 0.11136364 | PT r (Planum Temporale Right) |
| 1344 | 148 | -7.9336004 | 0.11011905 | IC r (Insular Cortex Right) |
| 2831 | 302 | -8.2489786 | 0.10667609 | SFG l (Superior Frontal Gyrus Left) |
| 409 | 43 | 4.26822662 | 0.10513447 | aMTG r (Middle Temporal Gyrus, anterior division Right) |
| 877 | 84 | -6.3631492 | 0.09578107 | CO r (Central Opercular Cortex Right) |
| 1235 | 118 | -6.0016136 | 0.09554656 | pSMG r (Supramarginal Gyrus, posterior division Right) |
| 3657 | 314 | -7.0040483 | 0.08586273 | PostCG l (Postcentral Gyrus Left) |
| 2593 | 213 | -7.4163237 | 0.08214423 | AC (Cingulate Gyrus, anterior division) |
| 5613 | 410 | -7.5889611 | 0.07304472 | Precuneous (Precuneous Cortex) |
| 2397 | 165 | -8.1614428 | 0.06883605 | PC (Cingulate Gyrus, posterior division) |
| 1357 | 84 | 4.52055645 | 0.06190125 | pMTG r (Middle Temporal Gyrus, posterior division Right) |
| 392 | 24 | -5.7097554 | 0.06122449 | pSTG l (Superior Temporal Gyrus, posterior division Left) |
| 1126 | 68 | 4.91622639 | 0.06039076 | SubCalC (Subcallosal Cortex) |
| 1064 | 62 | -5.9141626 | 0.05827068 | pSMG l (Supramarginal Gyrus, posterior division Left) |
| 2740 | 158 | -6.2265053 | 0.05766423 | MidFG r (Middle Frontal Gyrus Right) |
| 698 | 40 | 4.17123604 | 0.05730659 | Hippocampus r |
| 4813 | 240 | -6.8718615 | 0.04986495 | sLOC r (Lateral Occipital Cortex, superior division Right) |
| 766 | 37 | -5.5397215 | 0.04830287 | IFG oper l (Inferior Frontal Gyrus, pars opercularis Left) |
| 946 | 45 | -6.8813071 | 0.04756871 | pITG r (Inferior Temporal Gyrus, posterior division Right) |
| 641 | 28 | -5.4974313 | 0.04368175 | ICC l (Intracalcarine Cortex Left) |
| 2383 | 97 | 5.35161686 | 0.04070499 | TP r (Temporal Pole Right) |
| 519 | 21 | -5.839304 | 0.04046243 | Cuneal l (Cuneal Cortex Left) |
| 309 | 11 | -5.4827623 | 0.03559871 | HG l (Heschl's Gyrus Left) |
| 4967 | 157 | -6.335186 | 0.03160862 | sLOC l (Lateral Occipital Cortex, superior division Left) |
| 449 | 14 | 4.28958321 | 0.0311804 | aMTG l (Middle Temporal Gyrus, anterior division Left) |
| 2927 | 91 | -7.2173777 | 0.03108985 | MidFG l (Middle Frontal Gyrus Left) |
| 984 | 30 | 3.87097645 | 0.0304878 | MedFC (Frontal Medial Cortex) |
| 8081 | 197 | -6.0483851 | 0.02437817 | FP r (Frontal Pole Right) |
| 6926 | 142 | -6.8495755 | 0.02050245 | FP l (Frontal Pole Left) |
| 1732 | 33 | -6.0843434 | 0.01905312 | LG r (Lingual Gyrus Right) |
| 282 | 5 | -6.4777303 | 0.0177305 | HG r (Heschl's Gyrus Right) |
| 417 | 7 | -4.7737594 | 0.01678657 | pSTG r (Superior Temporal Gyrus, posterior division Right) |
| 867 | 11 | -5.3206539 | 0.01268743 | Putamen l |
| 686 | 8 | -4.9994578 | 0.01166181 | IFG oper r (Inferior Frontal Gyrus, pars opercularis Right) |

| **CMA ITS(+) vs ITS(-)** | | | | |
| --- | --- | --- | --- | --- |
| **ROI size (Voxel)** | **Cluster size (Voxels)** | **Max**  **(+/-) Value** | **Coverage** | **Region Label** |
| 643 | 457 | -7.3966374 | 0.71073095 | SMA L(Juxtapositional Lobule Cortex -formerly Supplementary Motor Cortex- Left) |
| 567 | 296 | -7.7636485 | 0.52204586 | PO l (Parietal Operculum Cortex Left) |
| 714 | 363 | -7.402133 | 0.50840336 | SMA r (Juxtapositional Lobule Cortex -formerly Supplementary Motor Cortex- Right) |
| 954 | 474 | -8.1466961 | 0.49685535 | aSMG l (Supramarginal Gyrus, anterior division Left) |
| 3233 | 1551 | -10.170628 | 0.47974018 | PostCG r (Postcentral Gyrus Right) |
| 877 | 389 | -8.3563004 | 0.44355758 | CO r (Central Opercular Cortex Right) |
| 1460 | 634 | -7.746273 | 0.43424658 | SPL l (Superior Parietal Lobule Left) |
| 309 | 134 | -6.6379743 | 0.43365696 | HG l (Heschl's Gyrus Left) |
| 3657 | 1546 | -8.7940531 | 0.42275089 | PostCG l (Postcentral Gyrus Left) |
| 440 | 175 | -7.3460622 | 0.39772727 | PT r (Planum Temporale Right) |
| 1476 | 564 | -8.0845442 | 0.38211382 | SPL r (Superior Parietal Lobule Right) |
| 565 | 209 | -8.1472616 | 0.3699115 | PT l (Planum Temporale Left) |
| 538 | 182 | -7.2837524 | 0.33828996 | PO r (Parietal Operculum Cortex Right) |
| 4275 | 1369 | -9.8609104 | 0.32023392 | PreCG r (Precentral Gyrus Right) |
| 801 | 250 | -8.0003824 | 0.31210986 | aSMG r (Supramarginal Gyrus, anterior division Right) |
| 946 | 273 | -7.2745805 | 0.28858351 | pITG r (Inferior Temporal Gyrus, posterior division Right) |
| 4359 | 1241 | -9.0350227 | 0.28469833 | PreCG l (Precentral Gyrus Left) |
| 1334 | 275 | -6.8304901 | 0.20614693 | IC l (Insular Cortex Left) |
| 1313 | 264 | -9.3622637 | 0.20106626 | PaCiG l (Paracingulate Gyrus Left) |
| 1361 | 268 | -6.7388096 | 0.19691403 | PaCiG r (Paracingulate Gyrus Right) |
| 2927 | 569 | -8.2537565 | 0.19439699 | MidFG l (Middle Frontal Gyrus Left) |
| 282 | 51 | -6.2008214 | 0.18085106 | HG r (Heschl's Gyrus Right) |
| 2831 | 502 | -8.3034325 | 0.1773225 | SFG l (Superior Frontal Gyrus Left) |
| 2740 | 482 | -7.1703086 | 0.17591241 | MidFG r (Middle Frontal Gyrus Right) |
| 766 | 123 | -6.5320768 | 0.16057441 | IFG oper l (Inferior Frontal Gyrus, pars opercularis Left) |
| 2397 | 360 | -7.4730868 | 0.15018773 | PC (Cingulate Gyrus, posterior division) |
| 978 | 144 | -6.298193 | 0.14723926 | CO l (Central Opercular Cortex Left) |
| 4967 | 644 | -6.8698282 | 0.12965573 | sLOC l (Lateral Occipital Cortex, superior division Left) |
| 1064 | 129 | -7.2672873 | 0.1212406 | pSMG l (Supramarginal Gyrus, posterior division Left) |
| 6926 | 767 | -8.2637014 | 0.11074213 | FP l (Frontal Pole Left) |
| 5613 | 603 | -7.569108 | 0.10742918 | Precuneous (Precuneous Cortex) |
| 2680 | 287 | -7.3181129 | 0.10708955 | SFG r (Superior Frontal Gyrus Right) |
| 2593 | 235 | -6.6857667 | 0.09062862 | AC (Cingulate Gyrus, anterior division) |
| 1732 | 151 | -5.2115149 | 0.08718245 | LG r (Lingual Gyrus Right) |
| 377 | 32 | -7.4157109 | 0.08488064 | PP r (Planum Polare Right) |
| 8081 | 603 | -7.1561079 | 0.07461948 | FP r (Frontal Pole Right) |
| 1344 | 94 | -5.9289389 | 0.06994048 | IC r (Insular Cortex Right) |
| 1235 | 86 | -6.3390174 | 0.06963563 | pSMG r (Supramarginal Gyrus, posterior division Right) |
| 1442 | 86 | -6.0123577 | 0.05963939 | FOrb r (Frontal Orbital Cortex Right) |
| 686 | 40 | -5.1570506 | 0.05830904 | IFG oper r (Inferior Frontal Gyrus, pars opercularis Right) |
| 4813 | 263 | -6.1250277 | 0.05464367 | sLOC r (Lateral Occipital Cortex, superior division Right) |
| 867 | 44 | -5.788403 | 0.05074971 | Putamen l |
| 278 | 14 | -5.222096 | 0.05035971 | aSTG r (Superior Temporal Gyrus, anterior division Right) |
| 719 | 30 | -5.3284492 | 0.04172462 | pTFusC r (Temporal Fusiform Cortex, posterior division Right) |
| 555 | 22 | -5.2122364 | 0.03963964 | IFG tri r (Inferior Frontal Gyrus, pars triangularis Right) |
| 519 | 20 | -5.6749573 | 0.03853565 | Cuneal l (Cuneal Cortex Left) |
| 392 | 15 | -6.1626425 | 0.03826531 | pSTG l (Superior Temporal Gyrus, posterior division Left) |
| 355 | 13 | -4.6176887 | 0.03661972 | FO l (Frontal Operculum Cortex Left) |
| 650 | 21 | -4.7522979 | 0.03230769 | IFG tri l (Inferior Frontal Gyrus, pars triangularis Left) |
| 1516 | 38 | -5.1194968 | 0.02506596 | LG l (Lingual Gyrus Left) |
| 536 | 13 | -5.2220273 | 0.02425373 | Caudate l |
| 342 | 7 | -4.672123 | 0.02046784 | Amygdala r |
| 1126 | 22 | -4.8422213 | 0.01953819 | SubCalC (Subcallosal Cortex) |
| 752 | 14 | -4.7688575 | 0.01861702 | ICC r (Intracalcarine Cortex Right) |
| 1680 | 31 | -5.6789756 | 0.01845238 | FOrb l (Frontal Orbital Cortex Left) |
| 1469 | 25 | -6.0837831 | 0.01701838 | AG r (Angular Gyrus Right) |
| 641 | 8 | -4.8663054 | 0.0124805 | ICC l (Intracalcarine Cortex Left) |
| 815 | 9 | -5.0860734 | 0.01104294 | TOFusC r (Temporal Occipital Fusiform Cortex Right) |

| **CMA PS(+) vs PS(-)** | | | | |
| --- | --- | --- | --- | --- |
| **ROI size (Voxel)** | **Cluster size (Voxels)** | **Max**  **(+/-) Value** | **Coverage** | **Region Label** |
| 567 | 392 | -8.4379187 | 0.69135802 | PO l (Parietal Operculum Cortex Left) |
| 643 | 440 | -10.065829 | 0.68429238 | SMA L(Juxtapositional Lobule Cortex -formerly Supplementary Motor Cortex- Left) |
| 714 | 406 | -9.4505987 | 0.56862745 | SMA r (Juxtapositional Lobule Cortex -formerly Supplementary Motor Cortex- Right) |
| 3657 | 2077 | -10.314614 | 0.56795187 | PostCG l (Postcentral Gyrus Left) |
| 954 | 512 | -7.9843431 | 0.53668763 | aSMG l (Supramarginal Gyrus, anterior division Left) |
| 877 | 460 | -8.9296198 | 0.52451539 | CO r (Central Opercular Cortex Right) |
| 3233 | 1691 | -9.3170605 | 0.52304361 | PostCG r (Postcentral Gyrus Right) |
| 538 | 236 | -8.7076845 | 0.43866171 | PO r (Parietal Operculum Cortex Right) |
| 309 | 134 | -7.0255322 | 0.43365696 | HG l (Heschl's Gyrus Left) |
| 565 | 227 | -7.9669709 | 0.40176991 | PT l (Planum Temporale Left) |
| 1460 | 581 | -8.5599499 | 0.39794521 | SPL l (Superior Parietal Lobule Left) |
| 440 | 173 | -7.429575 | 0.39318182 | PT r (Planum Temporale Right) |
| 355 | 123 | -6.4194212 | 0.34647887 | FO l (Frontal Operculum Cortex Left) |
| 1361 | 466 | -9.2316093 | 0.3423953 | PaCiG r (Paracingulate Gyrus Right) |
| 4275 | 1420 | -10.226765 | 0.33216374 | PreCG r (Precentral Gyrus Right) |
| 1334 | 435 | -9.3602219 | 0.32608696 | IC l (Insular Cortex Left) |
| 978 | 313 | -7.3821368 | 0.3200409 | CO l (Central Opercular Cortex Left) |
| 4359 | 1394 | -8.8621216 | 0.31979812 | PreCG l (Precentral Gyrus Left) |
| 801 | 244 | -7.5126333 | 0.30461923 | aSMG r (Supramarginal Gyrus, anterior division Right) |
| 1313 | 377 | -9.8989496 | 0.28712871 | PaCiG l (Paracingulate Gyrus Left) |
| 313 | 78 | -6.0821614 | 0.24920128 | FO r (Frontal Operculum Cortex Right) |
| 2740 | 635 | -11.081399 | 0.23175182 | MidFG r (Middle Frontal Gyrus Right) |
| 2927 | 638 | -8.0747318 | 0.21797062 | MidFG l (Middle Frontal Gyrus Left) |
| 2593 | 482 | -8.6916876 | 0.18588508 | AC (Cingulate Gyrus, anterior division) |
| 282 | 51 | -6.236927 | 0.18085106 | HG r (Heschl's Gyrus Right) |
| 2831 | 490 | -8.9317789 | 0.17308372 | SFG l (Superior Frontal Gyrus Left) |
| 766 | 122 | -7.0944505 | 0.15926893 | IFG oper l (Inferior Frontal Gyrus, pars opercularis Left) |
| 1476 | 233 | -6.7770209 | 0.15785908 | SPL r (Superior Parietal Lobule Right) |
| 686 | 108 | -7.5997715 | 0.1574344 | IFG oper r (Inferior Frontal Gyrus, pars opercularis Right) |
| 1344 | 169 | -6.6907072 | 0.12574405 | IC r (Insular Cortex Right) |
| 2680 | 322 | -8.2098017 | 0.12014925 | SFG r (Superior Frontal Gyrus Right) |
| 377 | 43 | -7.4166651 | 0.11405836 | PP r (Planum Polare Right) |
| 2397 | 249 | -8.8142252 | 0.10387985 | PC (Cingulate Gyrus, posterior division) |
| 8081 | 707 | -6.7181058 | 0.08748917 | FP r (Frontal Pole Right) |
| 946 | 81 | -6.1367745 | 0.08562368 | pITG r (Inferior Temporal Gyrus, posterior division Right) |
| 6926 | 588 | -7.6312928 | 0.08489749 | FP l (Frontal Pole Left) |
| 5613 | 457 | -8.0454731 | 0.08141814 | Precuneous (Precuneous Cortex) |
| 4967 | 367 | -6.5606804 | 0.07388766 | sLOC l (Lateral Occipital Cortex, superior division Left) |
| 1064 | 71 | -7.2163877 | 0.06672932 | pSMG l (Supramarginal Gyrus, posterior division Left) |
| 867 | 42 | -5.155375 | 0.04844291 | Putamen l |
| 521 | 21 | -5.905549 | 0.0403071 | Caudate r |
| 555 | 22 | -5.9494939 | 0.03963964 | IFG tri r (Inferior Frontal Gyrus, pars triangularis Right) |
| 519 | 20 | -5.6105666 | 0.03853565 | Cuneal l (Cuneal Cortex Left) |
| 641 | 22 | -5.3510809 | 0.03432137 | Cuneal r (Cuneal Cortex Right) |
| 1235 | 40 | -6.5025806 | 0.03238866 | pSMG r (Supramarginal Gyrus, posterior division Right) |
| 1732 | 50 | -5.4813409 | 0.02886836 | LG r (Lingual Gyrus Right) |
| 888 | 24 | -5.2013941 | 0.02702703 | OFusG r (Occipital Fusiform Gyrus Right) |
| 1680 | 42 | -6.2445183 | 0.025 | FOrb l (Frontal Orbital Cortex Left) |
| 4813 | 114 | -6.0733342 | 0.02368585 | sLOC r (Lateral Occipital Cortex, superior division Right) |
| 392 | 9 | -5.2772083 | 0.02295918 | pSTG l (Superior Temporal Gyrus, posterior division Left) |
| 278 | 6 | -5.6741691 | 0.02158273 | aSTG r (Superior Temporal Gyrus, anterior division Right) |
| 650 | 11 | -5.0651217 | 0.01692308 | IFG tri l (Inferior Frontal Gyrus, pars triangularis Left) |
| 73 | 1 | -4.7536492 | 0.01369863 | SCC l (Supracalcarine Cortex Left) |

| **SFA PS(+) vs PS (-)** | | | | |
| --- | --- | --- | --- | --- |
| **ROI size (Voxel)** | **Cluster size (Voxels)** | **Max**  **(+/-) Value** | **Coverage** | **Region Label** |
| 1460 | 393 | 6.50876331 | 0.26917808 | SPL l (Superior Parietal Lobule Left) |
| 269 | 62 | 7.84789944 | 0.23048327 | Pallidum r |
| 3657 | 645 | 7.70580435 | 0.17637408 | PostCG l (Postcentral Gyrus Left) |
| 2680 | 451 | 7.97844028 | 0.16828358 | SFG r (Superior Frontal Gyrus Right) |
| 302 | 47 | 6.77044153 | 0.15562914 | Pallidum l |
| 2831 | 415 | 8.5163002 | 0.14659131 | SFG l (Superior Frontal Gyrus Left) |
| 4359 | 527 | 10.0209751 | 0.12089929 | PreCG l (Precentral Gyrus Left) |
| 4275 | 469 | 7.6344018 | 0.1097076 | PreCG r (Precentral Gyrus Right) |
| 867 | 88 | 6.8408823 | 0.10149942 | Putamen l |
| 2042 | 203 | 9.20606613 | 0.09941234 | iLOC l (Lateral Occipital Cortex, inferior division Left) |
| 781 | 74 | 6.38528299 | 0.09475032 | toITG r (Inferior Temporal Gyrus, temporooccipital part Right) |
| 3233 | 306 | 9.64412689 | 0.09464893 | PostCG r (Postcentral Gyrus Right) |
| 946 | 83 | 5.97858953 | 0.08773784 | pITG r (Inferior Temporal Gyrus, posterior division Right) |
| 719 | 63 | 5.87644243 | 0.0876217 | pTFusC r (Temporal Fusiform Cortex, posterior division Right) |
| 954 | 83 | 7.36123753 | 0.0870021 | aSMG l (Supramarginal Gyrus, anterior division Left) |
| 2045 | 149 | 6.58297205 | 0.07286064 | iLOC r (Lateral Occipital Cortex, inferior division Right) |
| 815 | 53 | 6.28553057 | 0.06503067 | TOFusC r (Temporal Occipital Fusiform Cortex Right) |
| 804 | 49 | 7.29266214 | 0.06094527 | Putamen r |
| 801 | 48 | 6.59149265 | 0.05992509 | aSMG r (Supramarginal Gyrus, anterior division Right) |
| 1476 | 86 | 6.39809227 | 0.05826558 | SPL r (Superior Parietal Lobule Right) |
| 860 | 48 | 5.95039606 | 0.05581395 | pTFusC l (Temporal Fusiform Cortex, posterior division Left) |
| 6926 | 377 | 7.06787395 | 0.05443257 | FP l (Frontal Pole Left) |
| 698 | 37 | 6.79447699 | 0.0530086 | toITG l (Inferior Temporal Gyrus, temporooccipital part Left) |
| 2740 | 142 | 8.63149071 | 0.05182482 | MidFG r (Middle Frontal Gyrus Right) |
| 4967 | 241 | 6.49850941 | 0.04852023 | sLOC l (Lateral Occipital Cortex, superior division Left) |
| 652 | 28 | 6.62032175 | 0.04294479 | TOFusC l (Temporal Occipital Fusiform Cortex Left) |
| 714 | 30 | 5.83018494 | 0.04201681 | SMA r (Juxtapositional Lobule Cortex -formerly Supplementary Motor Cortex- Right) |
| 2927 | 115 | 9.65306759 | 0.03928937 | MidFG l (Middle Frontal Gyrus Left) |
| 519 | 19 | 5.43829775 | 0.03660886 | Cuneal l (Cuneal Cortex Left) |
| 5613 | 186 | 7.22500563 | 0.03313736 | Precuneous (Precuneous Cortex) |
| 326 | 10 | 5.07212114 | 0.03067485 | aITG r (Inferior Temporal Gyrus, anterior division Right) |
| 143 | 4 | 4.66062689 | 0.02797203 | SCC r (Supracalcarine Cortex Right) |
| 1442 | 38 | 6.30349064 | 0.02635229 | FOrb r (Frontal Orbital Cortex Right) |
| 8081 | 206 | 6.58714867 | 0.02549189 | FP r (Frontal Pole Right) |
| 888 | 21 | 5.23293066 | 0.02364865 | OFusG r (Occipital Fusiform Gyrus Right) |
| 4813 | 108 | 6.47951078 | 0.02243923 | sLOC r (Lateral Occipital Cortex, superior division Right) |
| 292 | 5 | 4.95111799 | 0.01712329 | aTFusC r (Temporal Fusiform Cortex, anterior division Right) |
| 521 | 8 | 5.34865952 | 0.01535509 | Caudate r |
| 752 | 11 | 5.10360146 | 0.01462766 | ICC r (Intracalcarine Cortex Right) |
| 1313 | 19 | 5.86572027 | 0.01447068 | PaCiG l (Paracingulate Gyrus Left) |
| 1516 | 19 | 5.55524254 | 0.01253298 | LG l (Lingual Gyrus Left) |
